# Heart-lung connections: Phenotypic and genetic insights from a large-scale genome-wide cross-trait analysis

**DOI:** 10.64898/2026.03.15.24309272

**Authors:** Jun Qiao, Yaixin Yao, Shiyu Zheng, Minjing Chang, Pengwei Zhang, Yuqing Yang, Yuxiao Kong, Yao Qiao, Jinyi Chen, Ziyi Han, Jialin Hou, Ning Tan, Ling Wang, Pengcheng He, Guo-chang Fan, Sakthivel Sadayappan, Anil G. Jegga, Wei Huang, Lei Jiang, Yuliang Feng

## Abstract

**Background:** Extensive comorbidity between cardiovascular (CVD) and respiratory (RT) diseases is well-documented, yet the shared genetic mechanisms remain elusive. Genetic pleiotropy may play a pivotal role in understanding the intricate comorbidity patterns associated with cardiovascular and respiratory conditions.

**Methods:** Our comprehensive analysis encompasses the largest available GWAS dataset of European ancestry covering six major CVDs (atrial fibrillation, coronary artery disease, venous thromboembolism, heart failure, peripheral arterial disease, and stroke) and four prevalent RTs (asthma, chronic obstructive pulmonary disease, idiopathic pulmonary fibrosis, and sleep apnea). Initially, we aimed to unveil the common genetic basis of major CVDs, through genome-wide and local genetic correlations and polygenic overlap. Subsequently, the shared genetic mechanisms between RTs and CVDs was investigated in terms of both horizontal and vertical pleiotropy. From a horizontal pleiotropy perspective, cross-trait analysis was utilized to identify pleiotropic genetic determinants including genomic loci, single nucleotide polymorphisms (SNPs), genes, biological pathways, and protein targets. From a vertical pleiotropic perspective, Mendelian randomization was employed to evaluate potential causal relationships between CVDs and RTs.

**Results:** Our study confirmed the significant existence of genetic correlations and overlaps between CVDs and RTs. Pleiotropy analysis under the composite null hypothesis identified 17,964 significant potential pleiotropic SNPs in 24 trait pairs, with 73 pleiotropic loci and 69 colocalized loci detected. Gene-based analysis revealed 59 candidate pleiotropic genes, highly enriched in unsaturated fatty acid biosynthetic processes and MHC class I-mediated antigen processing and presentation. Mendelian randomization analysis demonstrated a positive causal relationship only between chronic obstructive pulmonary disease and heart failure. Overall, the genetic basis between CVDs and RTs was inconsistent with vertical pleiotropy, suggesting the dramatic impact of horizontal pleiotropy.

**Conclusions:** Our findings indicate widely distributed pleiotropic genetic determinants between RTs and CVDs across the genome. These results support a common genetic basis for RTs and CVDs and are important for intervention and therapeutic targets in comorbidities.

**Clinical Perspective:** *What Is New?:*

1. A common genetic underpinning for CVDs and RTs has been identified using a variety of approaches and further explained as a shared genetic mechanism mediated by pleiotropy.
2. The systematic atlas of horizontal pleiotropy addressed key questions about pleiotropic SNPs, genomic loci, genes, functional features, and protein targets contributing to comorbidity between CVDs and RTs.
3. The systematic atlas of vertical pleiotropy highlighted causal associations between CVDs and RTs beyond the observed correlations.

*What Are the Clinical Implications?:* This study may help to elucidate the shared genetic mechanism between respiratory and cardiovascular diseases and further prioritize shared drug targets between RTs and CVDs.

## Introduction

Cardiovascular and respiratory diseases, collectively accounting for the highest global mortality rates, represent the foremost contributors to the worldwide burden of disease^1^. Cardiovascular diseases (CVDs) comprise a spectrum of heart and blood vessel illnesses brought on by the interplay of genetic, environmental, and lifestyle factors^2^. In contrast, chronic respiratory diseases constitute a multifaceted spectrum of airway or lung pathologies, characterized by inflammatory states such as chronic bronchitis, mucus overproduction, and repetitive injury to the alveolar epithelium^3-6^. Owing to the distinct organs and manifestations intrinsic to these categories, they have traditionally been subject to independent diagnoses, treatments, and research efforts. However, decades-long observational studies have underscored the strong correlation between increased CVD incidence and RTs. Maclagan *et al*. demonstrated that the most typical chronic respiratory disease, that is chronic obstructive pulmonary disease (COPD), exhibits a 25% higher incidence of major adverse cardiovascular events compared to patients without COPD^7^. The widespread co-morbidity of RTs and CVDs implies the existence of complex co-pathogenic patterns.

A plausible link may be genetic pleiotropy, a principle genetic mechanism wherein genetic variants affect multiple complex phenotypic traits by vertical and horizontal pleiotropy^8-10^. Horizontal pleiotropy arises when two phenotypes are affected by the same genetic variation, elucidating possible shared biological pathways linking complex traits. However, vertical pleiotropy arises when genetic variation influences one phenotype, the experience/expression of which then causes a second phenotype, this is the type of Mendelian randomization (MR) mainly targets. Genome-wide association studies (GWAS) have pinpointed thousands of genomic risk loci across multiple phenotypes, revealing a remarkable interconnectedness between RTs and CVDs^11-19^. Nevertheless, the shared genetic components and horizontal pleiotropics between RTs and CVDs remain elusive. Previous studies have primarily focused on explaining the genetic association of a single respiratory disease (e.g. COPD or asthma) with some CVDs^20, 21^, thus, they were unable to provide a systematic and complete view of the genetic architectures between the RTs and CVDs.

In this work, we encompass the largest available GWAS datasets in individuals of European ancestry of four widespread RTs (including asthma, chronic obstructive pulmonary disease [COPD], idiopathic pulmonary fibrosis [IPF], sleep apnoea [OSA]) and six major CVDs (including atrial fibrillation [AF], coronary artery disease [CAD], venous thromboembolism [VTE], heart failure [HF], peripheral artery disease [PAD] and stroke) and provide a comprehensive and innovative interpretation of the shared genetic architectures underlying the RTs and CVDs. First, shared genetic architectures between RTs and CVDs were evaluated by genome-wide and local genetic correlations and polygenic overlap. Second, a cross-trait analysis was applied to indicate shared genetic signals at the SNP and genomic loci level. Third, to explicate the credible influence mechanism of pleiotropic loci, dual identification methods (position mapping and expression quantitative trait loci [eQTL] mapping) were employed to identify genes related to pleiotropic loci and further explore their shared biological pathways. Fourth, causal plasma proteins and their possible targeted drugs were identified to facilitate potential clinical applications of comorbidities. Finally, we systemically evaluated the potential causal relationships of RTs and CVDs by Mendelian randomization analysis and sensitivity analysis.

## Materials and methods

### Data selection and quality control

In this study, GWAS summary statistics from European ancestry populations for four RTs and six CVDs were utilized. Detailed information about these diseases and their publication sources is available in Supplementary Table 1. Pre-analysis quality control steps were performed on GWAS summary statistics, including aligning with 1000 Genomes Project v3 Europeans reference of hg19 genome build, excluding non-biallelic SNPs, removing SNPs with duplicated or missing rs ID, and deleting SNPs with minor allele frequency (MAF) < 0.01. To ensure fair and interpretable comparisons between RTs and CVDs, only SNPs available for all these ten diseases (5,452,339 SNPs) were considered. Additional data processing operations were carried out in subsequent studies by the respective needs of different methodologies.

### Genome-wide genetic correlations by LDSC

We estimated genome-wide genetic correlations between CVDs and RTs using Linkage Disequilibrium Score regression (LDSC)^22^. LDSC (version 1.0.1), an effective tool for analyzing genetic correlations of complex diseases or traits^23^, utilized the linkage disequilibrium structure of the genome to estimate the distribution of individual SNP effect sizes to their LD scores. Pre-estimated LD scores were obtained from the 1000 Genomes Phase 3 genotyping data from European samples^24^. Due to its complex structure, the major histocompatibility complex (MHC) region (CHR6: 26-34 Mb) was excluded. First, univariate LDSC analysis was used to estimate the overall disease SNP-based heritability (h2SNP, i.e., the proportion of common genetic variation included in the study that explains phenotypic variation in the variable). Bivariate LDSC was then employed to estimate genetic correlations (rgs, i.e., a key population parameter that describes the common genetic structure of complex traits and diseases) between RTs and CVDs. Genetic correlations range from -1 to +1, with the sign indicating the same genetic factors contribute to variance in the target traits in opposite or same directions, respectively. *P*-values of the genome-wide genetic correlations below the Bonferroni-corrected threshold (*P* = 0.05 / 24 combinations = 2.08×10^-3^) were considered significant. LDSC applied to specifically expressed genes (LDSC-SEG) was used to identify enrichments in tissue-specific gene expression, with several gene sets including multi-tissue gene expression (including Genotype-Tissue Expression [GTEx] and Franke lab data)^25^. We include annotations constructed based on RNA sequencing data from 49 tissues from GTEx^24^ for tissue-specific gene expression, with Bonferroni-corrected significant threshold set at *P* < 0.05 / 49 / 10 traits = 1.02×10^-4^.

### Genome-wide polygenic overlap by MiXeR

We used univariate Gaussian causal mixture modeling method (MiXeR) analysis to quantify the polygenic overlap between RTs and CVDs. MiXeR (version 1.3)^26,27^, a statistical tool for analyzing genetic overlap between complex diseases or traits, estimated the total number of shared and trait-specific causal variants to quantify polygenic overlap irrespective of genetic correlation between traits. The 1000 Genomes Phase 3 genotyping data from European samples was used as a reference panel and excluded structurally complex MHC regions (CHR 6: 26-35 Mb)^24^. First, we applied univariate causal mixed models to calculate polygenicity (the proportion of non-null variants) and discoverability (the phenotypic variance of non-null variant effect sizes) for each trait. A bivariate causal mixed model was then constructed to estimate the total number of shared and phenotype-specific causal variants. In the cross-trait analysis, three scenarios were considered for each SNP and two traits: i) SNP affects both traits (shared SNP); ii) SNP affects only one of the two traits (trait-specific SNP); iii) SNP has no effect on either trait (null SNP). Bivariate MiXeR analysis modeled the additive genetic association of 2 traits as a mixture of 4 bivariate Gaussian components based on these assumptions. It determined the amount of causal variation unique and shared in traits between the two, presenting results in a Venn diagram. Following parameter fitting, the Dice coefficient, the ratio of shared variants to the total number of variants, was calculated to evaluate polygenic overlap on the 0-1 scale. Finally, we assessed model fit by comparing MiXeR model predictions to real GWAS data using Q-Q plots and the Akaike Information Criterion (AIC). A closer alignment of the model-based Q-Q plot to the actual Q-Q plot suggests more precise estimations by MiXeR. Positive AIC values support the MiXeR model of polygene overlap, indicating that GWAS data have sufficient power to distinguish overlap estimated by MiXeR from constrained models with minimum and maximum polygene overlap.

### Local genetic correlations by LAVA

The LDSC-calculated genome-wide genetic correlations represent the average correlation of genetic effects across the entire genome, implying that the contrast’s local r_g_s may nullify each other. To address this limitation, we employed the Local Analysis of [co]Variant Association (LAVA)^28^ to estimate local r_g_s between RTs and CVDs within specific genomic regions. LAVA estimated local r_g_s across 2495 regions of approximately equal size (∼1Mb) between two phenotypes, thus capable of identifying mixed effect directions despite minimal r_g_s^28^. The 1000 Genomes Phase 3 genotyping data from European samples were used as the LD reference panel following the procedure described in the original publication^24^. First, univariate analysis was used to evaluate local heritability for each trait. To enhance detection, a significant threshold of *P* < 1×10^-4^ was applied to filter out irrelevant regions. After identifying regions with sufficient univariate signal, 4,205 bivariate tests were conducted to evaluate pairwise r_g_s across pre-defined genomic regions. *P*-values of the local genetic correlations below the Bonferroni-corrected threshold (*P* = 0.05 / 4,205 combinations = 1.19×10^-5^) were considered significant.

### Cross-trait analysis by PLACO

The shared genetic basis can be interpreted as genetic variants impacting either both two traits (i.e. horizontal pleiotropy) or one trait through its influence on another trait (i.e. vertical pleiotropy)^8^. Therefore, to determine evidence for horizontal pleiotropy between RTs and CVDs, we next carried out a SNP-level pleiotropic test using pleiotropy analysis under composite null hypothesis (PLACO)^29^. PLACO was a novel statistical method for identifying pleiotropic loci between two traits by testing the composite null hypothesis that a locus is associated with zero or one of the traits. Only GWAS summary statistics was required as input, and the test statistic was created by multiplying the *Z* scores for the SNP in each of the two studies with the assumption that they follow a mixed distribution. SNPs with Z^2^ greater than 100 were excluded, due to the extremely large significance that may disproportionately influence the analysis.^22^ SNPs with *P*_PLACO_ < 5×10^−8^ were declared as significant pleiotropic variants.

### Genomic loci definition and functional annotation by FUMA

We defined independent genomic loci based on the functional mapping and annotation of genetic associations (FUMA, version 1.3), an online platform for SNPs and gene functional annotation^30^. We performed SNP2GENE to identify independently significant SNPs based on LD information from the 1000 Genomes Phase 3 genotyping data from European samples^24^. Before annotation, FUMA first defined independent significant SNPs that had a genome-wide significant *P*-value (5 × 10^-8^) and were independent at r2 < 0.6 within 1 Mb. Based on LD information, a subset of these independent significant SNPs was labeled as lead SNPs (independent from each other at r2 < 0.1). The gene annotations for all loci are determined by their proximity to the lead SNPs. The genomic loci were identified by merging the LD blocks of lead SNPs that are near each other (less than 500 kb apart). The top lead SNP was defined as the SNP with the lowest *P*-value in a specific region. We utilized LocusZoom^31^ to generate regional association plots with gene tracks, which allowed us to look at the details of each detected locus’s association. The directional effects of RTs and CVDs disease traits were assessed by comparing the z-scores of shared loci between them. Annotation was subsequently performed using the combined annotation dependent depletion (CADD) score, probability of regulatory functionality (RegulomeDB score), and transcription/regulatory effects from chromatin states (the minimum chromatin state)^32-34^. In detail, the CADD score is a score of SNP deleteriousness predicted by 63 functional annotations. SNPs with a CADD score > 12.37 were considered deleterious^32^. The RegulomeDB score is a categorical score that demonstrates the regulatory function of SNPs based on the overlap of available functional data, such as assignment to cis-expression quantitative trait loci (cis-eQTLs) and proof of transcription factor binding^33^. The accessibility of genomic areas is shown by the chromatin state, which is based on 15 category states predicted by ChromHMM from 5 chromatin marks for 127 epigenomes^34, 35^. We also used two gene mapping strategies in FUMA to map SNPs to genes for each disorder, including (1) positional mapping by physical position using a 1Mb window and (2) expression quantitative trait locus (eQTL) association.

We performed Bayesian colocalization analysis using the R package “coloc”^36^ for the FUMA-annotated pleiotropic loci to further identify potentially shared causal variants for the corresponding paired traits in each pleiotropic locus. Colocalization is a method to assess the presence of a shared causal variant in the region for two traits. The basic hypothesis for colocalization in the same genomic location is: (1) H0: neither trait has a causal genetic variant; (2) H1: only trait 1 has a causal genetic variant; (3) H2: only trait 2 has a causal genetic variant; (4) H3: both traits have a causal genetic variant, but not the same variant; (5) H4: both traits share the same causal variant. We declared genomic loci with PPH4 > 0.7 as colocalized loci with potentially shared causal variants. In addition, the SNP with the largest PPH4 in this locus will be identified as a candidate causal variant.

### Gene-level analysis by MAGMA, eMAGMA and TWAS

To further identify candidate pleiotropic genes, we performed performed gene-based association analysis using multi-marker analysis of genomic annotation (MAGMA)^37^ analysis based on PLACO and single-trait GWAS results. MAGMA yields gene-based *P*-values through the evaluation of the joint association effect of all SNPs within a gene while accounting for LD between SNPs^38^. The SNP locations were defined in reference to the human genome Build 37 (GRCh37/hg19), and only protein-coding genes (17,363) were included in the analysis. Then, we specified the boundaries of the gene length within the range of ‘10 kb outside the gene’, consistent with the default setting of position mapping using FUMA. *P*-values of the MAGMA analysis based on PLACO results below the Bonferroni-corrected threshold (*P* = 0.05 / 17,636 / 24 combinations = 1.18×10^-7^) were considered significant.

Conventional MAGMA primarily associates SNPs with genes based on physical proximity. However, mapping trait-associated SNPs to their nearest gene frequently fails to identify the functional gene because regulatory effects on gene expression can be long-range^39^. To remedy this shortcoming and improve the annotation and interpretation of GWAS association signals, we employed the eQTL multi-marker analysis of genomic annotation (eMAGMA)^40^ to systemically map genes using precomputed tissue-specific eQTL statistics based on PLACO results. eMAGMA, operating within the same statistical framework as MAGMA, transforms genome-wide association summary statistics into gene-level statistics by assigning risk variants to putative genes based on tissue-specific eQTL information rather than genomic positions^41^. To conduct this analysis, we utilized data from the Genotype-Tissue Expression Project (GTEx) v8, incorporating pre-computed gene expression weights from a comprehensive set of 49 tissues. However, considering the potential introduction of nuisance information when utilizing all tissues, we settled on analyzing a subset of 13 tissues informed by the outcomes of LDSC-SEG and insights from previous studies, ensuring a focused exploration of tissue-specific genes. In our meticulous investigation, an exhaustive set of tissue-specific genes was considered, encompassing 9,537 Adipose Subcutaneous-specific genes, 7,523 Adipose Visceral Omentum-specific genes, 7,833 Artery Aorta-specific genes, 3,494 Artery Coronary-specific genes, 9,546 Artery Tibial-specific genes, 2,718 EBV-Transformed Lymphocytes Cells-specific genes, 6,751 Heart Atrial Appendage-specific genes, 6,280 Heart Left Ventricle-specific genes, 3,223 Liver-specific genes, 8,345 Lung-specific genes, 9,497 Not Sun Exposed Suprapubic Skin-specific genes, 10,350 Sun Exposed Lower Leg Skin-specific genes and 8,245 Whole Blood-specific genes. The 1000 Genomes Phase 3 genotyping data from European samples was used to account for LD between SNPs. The significance threshold of eMAGMA associations was corrected with the Bonferroni correction (such as *P*_Adipose Subcutaneous_ < 0.05 / 9,537 / 24 combinations = 2.18×10^-7^, and *P*_Adipose Visceral Omentum_ < 0.05 / 7,523 / 24 combinations = 2.77×10^-7^).

Simultaneously, we also utilized the transcriptome-wide association study (TWAS)^42^ based on GWAS results to identify tissue-specific genes. TWAS integrates GWAS data with expression quantitative trait loci (eQTL) research to identify tissue-specific gene-trait correlations^43^, which has shown great promise both in interpreting GWAS findings and in elucidating the underlying disease mechanisms. Functional Summary-based Imputation (FUSION) was utilized to infer risk genes associated with RTs and CVDs using the GWAS summary statistics and gene expression weights while taking LD structures into account. The cis-genetic components of tissue-specific gene expression were computed by modeling all cis-SNPs using Best Linear Unbiased Prediction (BLUP), modeling SNPs and effect sizes (BSLMM), LASSO, Elastic Net and TOP SNPs. After cross-validating each desired model simultaneously, a final estimate of the weight of each desired model is obtained, and the gene expression weight selected was based on the prediction model with the best performance. Non-protein coding genes and genes with duplicated names were also removed for each tissue. We obtained tissue-specific *p*-values for each gene across different tissues and further performed Bonferroni correction for each tissue and selected significant genes.

### Pathway-level analysis by MAGMA and ToppFun

To explore the potential genetic pathways contributing to the comorbidity of RTs and CVDs, we performed MAGMA gene-set analysis. Gene-set tests in MAGMA were performed using a “competitive” approach whereby the test statistics for all genes within a particular gene-set (e.g. a biological pathway) were combined to obtain a joint association statistic. Gene-sets were defined from Gene Ontology (GO) biological processes and Reactome Knowledgebase using the Molecular Signatures Database (MsigDB). Gene definitions and their respective association signals for genes contributing to gene-sets were taken from the MAGMA gene-based analyses to identify potential biological processes that may be influenced by these markers. Multiple testing was accounted for by applying the Bonferroni-corrected threshold (*p* = 0.05 / (7,744 + 1,654) / 24 = 2.2×10^-7^). We then conducted gene set functional enrichment analysis for the genes that were significantly identified by MAGMA and eMAGMA analysis. Specifically, we performed a functional enrichment analysis using the mapped genes by the ToppFun (transcriptome, ontology, phenotype, proteome, and pharmacome annotations-based gene list functional enrichment analysis) function of ToppGene (https://toppgene.cchmc.org/), a well-known online software that detected the functional enrichment of a gene list based on functional annotations and protein interactions network. *p* < 0.05 was set as the threshold value^44^.

### Protein-level analysis by SMR and Colocalization analysis

To elucidate the potential association between plasma protein expression and the risk of diseases, we integrated summary statistics from plasma protein pQTLs and GWAS summary statistics for diseases using Summary data-based Mendelian Randomization (SMR)^35^. The pQTL data were sourced from a large-scale study of protein quantitative trait loci involving 35,559 Icelanders^45^, providing summary-level statistics of genetic associations with 4,907 plasma protein levels. For each protein, Cis-acting SNPs within ±1 Mb from the target gene’s transcription start site were considered. In this study, Only proteins with cis-pQTLs available at the genome-wide significance level (*p* < 5 × 10^-8^) were included in the SMR and colocalization analysis. As an extension of the MR, SMR (v.103) was developed to estimate pleiotropic associations between genetically determined traits and complex traits of interest using genome-wide significant SNPs as instrumental variables. Due to the presence of a shared and potentially causal variant at a locus, we performed a heterogeneity of associated instrumentation (HEIDI) test to explore whether there was a linkage in the observed associations. A *P*_HEIDI_ < 0.01 indicated that the observed connection may be caused by two different genetic variants that are in strong linkage disequilibrium with one another. Additionally, due to the limitation of using a single variant as the genetic instrument in SMR, which makes it challenging to discern horizontal pleiotropy or causality, we implemented the sensitivity test. This involved using summary statistics from multiple SNPs (multi-SNPs SMR)^46^ with a significance level of *P*_smr-muli_ < 0.05 to strengthen the primary analysis. To correct for multiple testing, false discovery rate (FDR) at a = 0.05 based on the Benjamini-Hochberg method was applied. Subsequently, we also employed the ‘coloc’ R package for genetic colocalization analysis to investigate whether the identified associations between proteins and diseases were driven by the same causal variant or linkage disequilibrium. Strong evidence of colocalization between GWAS and pQTL exists if the posterior probability of the shared causal variant (PP.H4) is > 0.7.

To assess the druggability potential of plasma proteins shared by RTs and CVDs, we queried OpenTargets^47^ public data for identified proteins. The Open Targets platform provides target-centric or disease-centric workflows for drug target identification and prioritization, incorporating data on the relationship between targets and diseases from various sources. The drugs associated with the target proteins in our study were categorized according to the clinical trial stage reported on the clinical trial website. Furthermore, we cross-referenced our results with lists of druggable genes to ensure consistency and replicability of our findings.

### Mendelian Randomization Analysis using LHC-MR and LCV

In our quest for more accurate and effective causal associations between RTs and CVDs, we employed the Latent Heritable Confounder MR (LHC-MR)^48^ approach. LHC-MR utilizes GWAS summary statistics and genome-wide genetic markers to estimate bidirectional causal effects, direct heritability effects, and confounding effects while accounting for sample overlap. This framework allows LHC-MR to distinguish SNPs based on their common association with a pair of traits and to differentiate heritable confounding factors leading to genetic associations from true causal relationships. Here unbiased bidirectional causal effects between the traits of RTs and CVDs were estimated simultaneously with the confounding effects of each trait, ensuring a comprehensive analysis. For forward causality, significance was first determined by the Bonferroni-corrected threshold (*P* = 0.05 / 24 combinations = 2.08×10^-3^), while simultaneously excluding the reverse causal causality (*P* = 0.05 / 24 combinations = 2.08×10^-3^), and vice versa. Our sensitivity study encompassed conventional bidirectional MR models, including Inverse-variance weighted (IVW), MR Egger, Weighted median, Simple mode, Weighted mode^49, 50^, considering *P*-values less than 0.05 as significant.

To further quantify evidence of causation, we implemented a Latent Causal Variable (LCV)^51^ model to estimate the genetic causality proportion (GCP) between RTs and CVDs. LCV minimized false-positive results in the context of high genetic relatedness and large sample sizes. Based on GWAS summary statistics and LD scores, LCV utilized LDSC-estimated cross-trait genetic correlations to account for genetic correlations between the two traits and applied the mixed fourth moment of bivariate effect size distributions to derive the posterior mean and standard deviation estimates of the GCP. The GCP described the proportion of the genetic component of one trait affecting the other trait, where GCP > 0 implied partial genetic causality of trait one on trait two and vice versa. The weak GCP estimated close to zero for genetically associated variables suggested potential mediation by horizontal pleiotropy, indicating shared genes. An absolute value of GCP > 0.6 was considered strong evidence of partial genetic causation, with a Bonferroni-corrected significant threshold of *P* < 0.05 / 24 combinations = 2.08×10^-3^.

## Result

### Estimation of global genetic correlations between RTs and CVDs

First, we present the outcomes of our analysis, with a focus on SNP-based heritability (h^2^_snp_) and genetic correlations (r_g_s) between our RTs and six CVDs. For CVDs, SNP-based heritability was less than 1% in three diseases, including HF (h^2^_snp_ = 0.82%, standard error [se] = 0.00070), PAD (h^2^_snp_ = 0.96%, se = 0.00130) and Stroke (h^2^_snp_ = 0.61%, SE = 0.00050). Conversely, CAD (h^2^_snp_ = 3.23%, se = 0.00190) exhibited relatively higher heritability levels. Notably, respiratory disorder COPD (h^2^_snp_ = 1.82%, se = 0.00100) indicated the lowest heritability level still exceeding 1%, while IPF demonstrated remarkably high heritability at 21.3%. In the 24 bivariate genetic correlations calculated by bivariate LDSC, all the pairwise genetic correlations with a *P*-value below 2.08×10^-3^ (0.05/24) were positive, averaging 0.272. The individual correlation ranged from 0.105 (*P* = 2.48×10^-5^) between Asthma and AF to 0.546 (*P* = 3.77×10^-25^) between COPD and PAD. Notably, approximately 75% (18/24) of these cross-trait genetic association estimates remained significant even after Bonferroni correction (Supplementary Table 2).

### Estimation of total polygenic overlap between cardiovascular and respiratory diseases

Within the selected four respiratory diseases, univariate MiXeR revealed that OSA exhibited the highest estimated polygenicity values, with 8,273 (SE = 894) OSA-influencing variants predicted. This suggests a potential involvement of numerous causally related genomic regions in the polygenic architecture of the disease. Furthermore, HF demonstrated higher polygenic characteristics compared to the other five cardiovascular diseases, with 2,467 (SE = 263) influential variants. Bivariate MiXeR analysis unveiled a diverse range in the measure of genetic overlap between each respiratory diseases and cardiovascular diseases. Notably, a significant polygenic overlap emerged among the four respiratory diseases and six cardiovascular diseases, with the Dice coefficient ranging from 0.0035-0.511. The most substantial genetic overlap was observed between OSA and HF, with 2,093 (SE = 322) shared variants, accounting for 25.3% and 84.0% of variants influencing OSA and HF, respectively. Conversely, fewer shared variants were identified between Idiopathic Pulmonary Fibrosis (IPF) and Atrial Fibrillation (AF), with only 15 (SE = 8) shared variants at 22.2% and 4.08% for IPF and AF, respectively. Despite the range in disease correlation strength (r_g_s range: -0.05 - 0.52), both relatively high positive genetic correlation (COPD and HF) and unique negative genetic correlation (IPF and AF) were observed. Notably, the relationship pair (IPF and PAD) with a correlation test of 0 still has a certain degree of genetic overlap. This phenomenon may be attributed to a pattern of balance effects involving concordant and discordant variants among shared variants, leading to the minimal genetic correlation (Supplementary Table 3, Supplementary Figure 1).

### Estimation of cross-trait local genetic correlation between cardiovascular and respiratory diseases

Despite the valuable insights provided by genetic correlation, its limitation lies in measuring an average effect size across the entire genome^52^, which may overlook detailed patterns at local genetic loci. It occurs due to the potential dilution of the global estimate by a mixture positive and significantly negative genetic correlations^27,53^. Therefore, our subsequent focus shifted towards examining local genetic regions to uncover potential associations between CVDs and RTs. Following the application of a filter based on univariate heritability (p < 1×10^-4^, Supplementary Table 4), pairs of traits that passed the initial test proceeded to bivariate assessments. This resulted in a total of 4,205 bivariate tests, involving 93 and 350 regions per RTs-CVDs trait pairs. Notably, only local correlations deemed significant after the Bonferroni-corrected *P*-value threshold of *P* < 0.05/4205 were considered significant and subjected to interpretation. Across all trait combinations, we identified 17 significant bivariate local regional correlations (local r_g_s range: -1.000 - 0.995; *P*-value range: 5.97×10^-12^ - 1.11×10^-05^). These comprised four instances of negative local genetic correlation between phenotype pairs and 13 instances of positive correlation between trait pairs (Supplementary Table 5). Intriguingly, evidence of a negative association between IPF and AF was observed in both the global correlation test LDSC and the local correlation test LAVA. Although this negative association did not meet the strict Bonferroni test threshold in LDSC, the consistent observation in both analyses merits attention. When considered alongside the findings of a previous cohort study by Hubbard et al., which reported no increased risk of AF in IPF patients compared to other respiratory diseases^54^, it suggests that the relationship between IPF and AF may not be a straightforward complication and warrants further investigation.

### Identification and functional annotation of shared genetic loci between cardiovascular and respiratory diseases

In total, PLACO identified 17,964 SNPs associated with potential pleiotropic variants across 24 pairs of disorders. FUMA further pinpointed 556 lead pleiotropic SNPs, mapping to 461 independent genomic risk loci (Supplementary Figure 2). Notably, the pleiotropic regions 16q12.2 and 19q13.32 exhibited consistent associations across numerous pairwise traits, encompassing Asthma, OSA and cardiovascular disease-related traits pairs (Supplementary Figure 3). This underscores the broad pleiotropic effects and the significance of these regions. A mixed pattern of allelic association was observed in 461 loci annotated by FUMA as pleiotropic. In particular, 199 of the top 423 SNPs (43.2%) exhibited a concordant effect for both of the diseases within a pair of traits, suggesting a synchronous influence on reducing or increasing the risk of respiratory disease and cardiovascular disease. Conversely, the remaining 140 of the top 461 SNPs (56.8%) showed inconsistent associations, hinting at potentially opposing biological mechanism (Supplementary Table 6-8).

Among the 461 index SNPs representing shared effects risk locus, 24 (5.2%) were identified as exonic variants, comprising 19 exonic variants of mRNA and five non-coding variants of exonic RNA. Notably, 39 leads SNP had CADD scores above the 12.37 threshold, indicating high deleteriousness^32^. The highest CADD score was attributed to rs116843064 in 19p13.2. In addition, 6.9% of candidate SNPs (n = 32, RegulomeDB scores ≤ 2) were categorized as functional, likely affecting binding. Of particular interest is rs5743618 at 4p14, simultaneously identified in three asthma-involved pairs (HF, VTE, and Stroke) among 19 exon variants. Although previously associated with asthma, its newfound impact on these three cardiovascular diseases underscores the potential of its pleiotropic loci and nearby genes (Supplementary Table 6). Notably, *TLR1* located near rs5743618, participates in the innate immune response to microbial agents, suggesting a common biological mechanism shared between respiratory and cardiovascular systems.

In a subsequent colocalization analysis, 69 SNPs from corresponding loci were identified as candidate-shared causal variants in 73 of 461 possible pleiotropic loci (15.8%). Notably, the 3q22.3, 15q22.33 and 19q13.32 locus were concurrently identified in pleiotropic loci for 2 pairs of traits (Asthma-CAD and COPD-CAD) but colocalized only between Asthma and CAD. They shared the same potential causal variant rs12637102, rs56062135 and rs8108864. Similarly, the potential causal variant site rs1230666 (located at 1p13.2) was preliminarily identified in four pairs of traits (Asthma-CAD, COPD-CAD, IPF-CAD, OSA-CAD), but only the Asthma-CAD and IPF-CAD relationship pairs are supported by colocalization evidence (Supplementary Table 6, Supplementary Figure 4).

### Identification of pleiotropic susceptibility genes through position mapping

Two position-based strategies were employed to discern susceptibility genes with pleiotropic effects in diseases, with MAGMA outcomes substantiated verified by FUMA position mapping. First, we conducted MAGMA association analysis of potential pleiotropic genes located at 73 remarkable loci passing COLOC. Notably, 97 risk genes (Supplementary Table 9), spanning the genome-wide region, exhibited statistical significance (*P* < 0.05 / 17,636 / 24). In addition, comparing the results of MAGMA using GWAS data of respiratory diseases and cardiovascular diseases, it was observed that 31 genes (32%) related to respiratory diseases were newly defined as related to cardiovascular diseases. In addition, pleiotropic effects of 43 entirely novel genes (44.3%) were identified. Eleven genes were recurrently identified in two or more trait pairs. Intriguingly, *DMWD*, *FBXO46*, *RSBN1*, *RSPH6A*, and *PHTF1* were discerned in two trait pairs associated with CAD (Asthma-CAD and IPF-CAD), where *PHTF1* was newly identified in both Asthma and IPF, while others were unique for CAD. Secondly, a total of 1,627 potential susceptibility genes were obtained in positional mapping performed using the SNP2GENE function of FUMA (Supplementary Table 10-12). Notably, all 97 risk genes identified by MAGMA were corroborated in FUMA position mapping, underscoring the robustness of the results.

### Identification of tissue-specific pleiotropic susceptibility genes by eQTL mapping

To discern the tissues which these SNPs exert their effects, we employed publicly available GWAS results and genotype tissue expression data from GTEx. Utilizing the LDSC-SEG method, we identified SNP heritability enrichment of specific tissues and functional categories. After correcting for the number of tested functional categories within traits (*P* < 1.02×10^-4^), we found significant SNP heritability enrichment for AF-related SNP in the Heart’s Left Ventricle, and CAD-related in the Artery Tibial (Supplementary Table 13, Supplementary Figure 5). Previous studies by Valette K et al.^55^ and Zhou Y et al.^21^ demonstrated skin tissue (especially tibial skin) enrichment in Asthma and AF, suggesting possible pleiotropic effect of skin tissue on respiratory diseases and cardiovascular diseases, guiding our focus on skin tissue as a potential concentration sites of genes. Subsequently, we selected 13 tissues, including the heart and skin, for further investigation of tissue-specific pleiotropic genes.

Initially, SNPs were assigned to putative genes, further utilizing tissue-specific cis-eQTL information through eMAGMA method, enriched in the selected 13 tissues. Among these, 2,535 tissue-specific genes passed the normal test, of which a total of 1281 pleiotropic genes passed the Bonferroni test in at least one relevant tissue (Supplementary Table 14). Notably, *ATF6B* and *PRRT1* were duplicated in eight trait pairs (Asthma-six CVDs, COPD-CAD, and IPF-VTE), suggesting higher tissue specificity in five tissues (adipose subcutaneous, adipose visceral omentum, lung, suprapubic skin not exposed to the sun, and lower leg skin exposed to the sun). In addition, 10 genes (*AGER*, *ATP6V1G2*, *BAG6*, *GSDMB*, *HLA-C*, *HLA-DQB1*, *HLA-DRB1*, *ORMDL3*, *PSORS1C1*, and *PSORS1C2*) occurred multiple times in eight trait pairs. These pleiotropic genes per trait were further validated for tissue-specificity through TWAS analysis (Supplementary Table 14). Furthermore, leveraging GTEx8 datasets by FUMA, we mapped SNPs to genes with significant eQTL associations, identifying 1,862 eQTLs for 24 trait pairs within 13 different tissues. Notably, 858 of these candidate genes were physically located outside the GWAS risk loci (Supplemental Table 10).

In conclusion, a total of 59 pleiotropic genes were identified by combining position-based MAGMA and eQTL-based eMAGMA methods. Among them, 22 genes were identified that are associated with the respiratory and cardiovascular systems for the first time, presenting as potential targets for drugs development and therapeutics (Supplementary Table 15).

### Shared mechanisms between cardiovascular and respiratory diseases

We further performed Toppfun webtool to study the biological functional annotations of the overlapped 59 significant pleiotropic genes.A total of eleven important biological process pathways were identified, involving plasma phospholipid n-3 fatty acids, class I MHC mediated antigen processing, fatty acid desaturases, lung fibrosis, omega-6 fatty acid desaturase activity, omega3/omega6 fatty acid synthesis, metabolism ofalphalinolenic acid, omega 9 fatty acid synthesis, telomere cap complex, tumor necrosis factor superfamily and tall1 pathway.

In addition, MAGMA Gene set enrichment analysis identified 85 significantly enriched pathways (Bonferroni-corrected significant P < 2.2×10^-7^), including 81 GO terms and 4 Reactome Gene Sets pathways(Supplementary Table 16). There was again evidence of pathways that involved the regulation of compound metabolic process and specific immunity being enriched. For example, negative regulation of the nucleotide metabolic process (GO: 0045980) and positive regulation of RNA metabolic process (GO: 0051254) were identified in 4 pairs of traits. Significant enrichment was observed in three traits for negative regulation of the biosynthetic process (GO: 0009890) and positive regulation of the macromolecule biosynthetic process (GO: 0010557). Meanwhile alpha beta t cell activation (GO: 0046631) were identified in 3 pairs of asthma-related traits. Interestingly, specific immune pathways were newly enriched in cardiovascular diseases, indicating their potential to provide new insights into pathogenesis.

### Assessment of causal proteins and their druggability in cardiovascular and respiratory diseases

Multiple testing corrections were applied to control for genome-wide type I errors, demonstrating strong evidence of association (FDR < 0.05). Simultaneously, the the Heterogeneity in Dependent Instruments (HEIDI) test (PHEIDI > 0.01) was conducted to investigate whether associations were attributed to shared causal variants. Subsequently, colocalization analysis was performed, to rule out confounding by LD. With a threshold of PPH4 > considered strong evidence of co-localization between GWAS and pQTL for traits. A total of 25 association signals were identified for RTs loci and 50 association signals for CVDs loci. Notably, four proteins (TMEM106B, TNFSF12, WBP2 and EFEMP1) were shared by respiratory and cardiovascular diseases. Among them, TMEM106B, TNFSF12 and EFEMP1 showed consistent negative effects in RTs and CVDs, while WBP2 showed opposite effects in these conditions. Unfortunately, proteins associated with HF, PAD and OSA did not pass the screening criteria (Supplementary Table 17, Supplementary Figure 6, Supplementary Figure 7).

For the 75 causal proteins supported by evidence of colocalization analyses, we assessed protein-drug interactions based on the OpenTargets platform. Eleven proteins were predicted to be druggable, targeting 48 existing drugs in clinical phase I to IV. Notably, 12 of these drugs are in phase IV trials, making stem strong candidates for potential therapeutic options (Supplementary Table 18). Of particular interest, TNFSF12, identified by SMR as associated with both AF and Asthma, demonstrated potential efficacy in treating respiratory and cardiovascular disorders with its targeting antibodies Biib-023 and Ro-5458640. Despite 56 causal proteins in the OpenTargets database having no drug-related information, 31 displayed extremely reliable druggability tiers, rendering them attractive candidates for potential future treatments.

### Mendelian Randomization between cardiovascular and respiratory diseases

Using LHC-MR methods, we investigated bidirectional causation effects between cardiovascular and respiratory diseases (Supplementary Table 19). Six pairs of direct causal effects of respiratory diseases on cardiovascular diseases were identified, encompassing five pairs with negative effects (Asthma-AF, asthma-VTE, COPD-HF, OSA-PAD, OSA-stroke), along with positive effects of IPF on CAD evidence. In addition, evidence of a negative causal effect of PAD and stroke on Asthma was found, while three pairs (Asthma-HF, COPD-CAD, COPD-PAD) of bidirectional potential relationships were rejected. Several alternative Mendelian randomization methods produced results generally consistent with the main study. For instance, Asthma versus VTE, PAD versus Asthma, and COPD versus HF were all confirmed by Inverse Variance Weighting (IVW) to exhibit evidence of negative causal effects (Supplementary Table 20). We further constructed the LCV models to verify a partial causal relationship between each RTs and CVDs disorder (Supplementary Table 21). Notably, all three methods provide evidence for a partial genetic causal relationship between COPD and HF, indicating that COPD may partially cause HF.

## Discussion

Clinical and epidemiological studies suggested a co-occurrence of RTs and CVDs, yet the genetic underpinnings remained poorly understood. In the present study, utilizing the largest GWAS summary statistics for four RTs (Asthma, COPD, IPF, and OSA) and six CVDs (AF, CAD, VTE, HF, PAD, and Stroke), we conducted a comprehensive large-scale genome-wide cross-trait analysis. By delving into the SNP, gene, pathway, and protein levels, we systematically elucidated the shared genetic architecture of RTs and CVDs and their detailed genetic mechanisms.

To this end, we first used multiple approaches with different model assumptions to provide complementary evidence to delve into the genetic architectures between RTs and CVDs from different perspectives. In detail, we demonstrated extensive genome-wide and local genetic correlations and multigene overlap between RTs and CVDs using LDSC, LAVA and MixeR methods, respectively. LDSC showed significant genetic association estimates for 75% (18/24) of these crossover traits. LAVA detected 71% (17/24) of significant bivariate local genetic correlations. Mixer discovered a high degree of genetic commonality between CVDs and RTs. Notably, we identify locally shared genetic mechanisms that have been overlooked. Specifically, despite the absence of any indication in the LDSC results regarding a genetic correlation between IPF-PAD and IPF-CAD traits, MixeR identified considerable genetic overlap between these two trait pairs. Furthermore, the LAVA analysis, conducted with less stringent *P*-value thresholds, illuminated local negative genetic correlations and local positive genetic correlations between the IPF-CAD and IPF-PAD trait pairs. The interplay of positive and negative genetic correlations was pivotal in explaining this phenomenon, underscoring the robustness of our research methodology.

We then further investigated the shared genetic mechanisms between RTs and CVDs which may be driven by pleiotropy. Initially, we examine from the standpoint of horizontal pleiotropy. PLACO analysis was utilized to undertake pleiotropic analysis at the variant level to validate the basis for shared genetic risk across diseases, and a total of 17,964 pleiotropic SNPs were detected. We further implemented functional annotation to investigate potential pleiotropic regions between RTs and CVDs. Of the 461 independent genomic risk loci mapped, 43.2% of genes showed consistent effects on both diseases within a trait pair, and 56.8% showed discordant associations. Among them, the gene locus with the highest pleiotropy is *FTO*, located on 16q12.2, which encodes fat mass and obesity-associated protein and may be closely related to an increased risk of asthma. Experiments have shown that *FTO* knockout mice exhibit a strong asthma-like phenotype upon allergen challenge, which is due to defects in airway epithelial ciliated cells^56^. In our study, we found that the *FTO* gene is also related to CADs, HF, PAD, and VTE. Afterward, in addition to demonstrating high colocalization, we also focused on nonsynonymous SNPs that may be detrimental to protein structure and function. The variant with the highest CADD score was rs116843164 in the 19p13.2 locus, which is an exonic variant of *ANGPTL4*. *ANGPTL4* encodes a protein secreted by adipose tissue, namely Angiopoietin-like 4. ANGPTL4 protein is selectively expressed in the fibroblast area of IPF lungs^57^, and its expression is increased in epicardial adipose tissue (EAT) to affect the occurrence of CAD^58^. ANGPTL4 is critical in the progression of IPF and CAD and may be a potential therapeutic target for patients with co-existing IPF and CAD. RegulomeDB score showed that the variant rs5743618 located at the 4p14 site of TLR1 has a high regulatory level and affects Asthma, HF, Stroke, and VTE by participating in the innate immune response to microbial agents^59-61^.

In addition, the existence of pleiotropy was demonstrated in our results at the gene-level. 32% of genes related to respiratory diseases were newly defined as related to cardiovascular diseases, and 44.3% of completely novel genes were pleiotropic. Among them, *DMWD*, *FBXO46*, *RSBN1*, *RSPH6A,* and *PHTF1* genes were identified in both trait pairs Asthma-CAD and IPF-CAD. Shared genetic determinants also reflect common biological pathways, with the unsaturated fatty acid production process being highlighted in the Toppfun enrichment analysis results. Previous study has demonstrated that the arachidonic acid pathway is involved in the development of Asthma, COPD, IPF, and OSA^62-65^, as well as playing a major role in the progression of cardiovascular system diseases.

We conducted proteome-wide MR and colocalization analysis to investigate the causative involvement of 1,708 circulating proteins in respiratory and cardiovascular diseases to provide preclinical clues for medication development. Notable causal proteins in our findings are tumor necrosis factor ligand superfamily member 12 (TNFSF12), also known as TNF-related weak inducer of apoptosis (TWEAK), which has a negative causal role in both Asthma and AF via TWEAK/Fn14 axis^66^. Specifically, the TWEAK/Fn14 axis accelerates the proliferation and migration of human airway smooth muscle cells (HASMC) by activating the NF-κB pathway, thereby exacerbating asthma airway remodeling^67^. Moreover, the TWEAK/Fn14 axis can mediate the hypertrophy of HL-1 atrial myocytes through the activation of the JAK2/STAT3 pathway, thereby affecting the occurrence and development of atrial fibrillation^68^. TWEAK-targeting antibodies Biib-023 and Ro-5458640 may become effective treatments for Asthma and AF.

Finally, we attempted to elucidate whether a causal relationship exists between RTs and CVDs using multiple Mendelian randomization methods, the core of which is to use vertical pleiotropy to examine the existence of a causal relationship. LHCMR and LCV methods showed a significant causal relationship only between COPD and HF, consistent with the notion that most genetic correlations between these traits arise from horizontal pleiotropy. On the other hand, there are high genetic correlations between RTs and CVDs trait pairs, a situation that may generate too many false positives in MR. And in this case, LCV exhibits relatively well-calibrated Type 1 and Type 2 errors. It is further suggested that vertical pleiotropy, at least one causal direction, may not explain the well-known high coexistence rates between RTs and CVDs.

Despite these valuable insights, our study has limitations. Firstly, it focused on four RTs and six CVDs. As more datasets become available, a more comprehensive analysis encompassing a broader spectrum of diseases should be performed. Secondly, our characterization of pleiotropic common variants was limited to individuals of European-ancestry due to statistical power considerations, potentially limiting the generalizability of current findings. Future research on rare genetic variants and diverse ancestries will provide a more comprehensive understanding of shared biology across these disease classes. Lastly, in terms of causal inference, while common genetic effects predominantly explain our results, we acknowledge the possibility of cyclical feedback loops between phenotypes, possibly for every pair of phenotypes.

## Conclusion

In summary, our study unveils the intricate genetic relationships between RTs and CVDs, providing new insights into the genetic architecture distributed across the genome, including pleiotropic SNPs, genes, as well as biological pathways. Furthermore, our research offer valuable insights into potentially druggable proteins and causal relationships, presenting innovative perspectives for the prevention and treatment of RTs and CVDs,.

## Data availability

GWAS summary statistics on Asthma and COPD are available from the Global Biobank Meta-Analysis Initiative (GBMI). Genetic information for OSA was obtained from the FinnGen consortium website: https://www.finngen.fi/en. GWAS summary statistics on IPF was obtained from International IPF Genetics Consortium. Genome-wide summary statistics on AF, HF, and Stroke are available at the GWAS Catalog (GCST90104539, GCST009541, and GCST90104539). GWAS summary statistics on CAD and PAD are publicly available for download at the Cardiovascular Disease Knowledge Portal (CVDKP) website: https://cvd.hugeamp.org/datasets.html. Genome-wide summary statistics on VTE are obtained from the deCODE genetics website: https://www.decode.com/summarydata/.

## Code availability

All software (and version, where applicable) used to conduct the analyses in this paper are freely available online:

LDSC (v1.0.1; https://github.com/bulik/ldsc), MiXeR (v1.3; https://github.com/precimed/mixer), LAVA (v0.1.0; https://github.com/josefin-werme/LAVA), FUMA (v1.5.4; http://fuma.ctglab.nl/), MAGMA (v.1.08; https://ctg.cncr.nl/software/magma), TWAS (http://gusevlab.org/projects/fusion/), SMR (v1.31; https://yanglab.westlake.edu.cn/software/smr/), COLOC (v5.2.1; https://github.com/chr1swallace/coloc), R (v.4.1.3; https://www.r-project.org/), LHC-MR (v0.0.0.9000; https://github.com/LizaDarrous/lhcMR), and LCV (https://github.com/lukejoconnor/LCV).

## Sources of Funding

This study was supported by Shenzhen Pengcheng Peacock Plan (To Y.F), National Natural Science Foundation (Grant no. 82170339 and 82270241), NSFC Incubation Project of Guangdong Provincial People’s Hospital (Grant no. KY0120220021), Natural Science Foundation of Guangdong Province (Grant no. 2023B1515020082) (To L.J.), National Institutes of Health R01 (Grant no. HL163148)(To W.H.) and Center for Computational Science and Engineering at Southern University of Science and Technology. The funder had no role in the design, implementation, analysis, interpretation of the data, approval of the manuscript, and decision to submit the manuscript for publication.

## Acknowledgements

J.Q., Y. F., L.J., and W.H. conceptualized, supervised this project and wrote the manuscript. J.Q., Y.Q., and J.C. performed the main analyses and wrote the manuscript. J.Q., M.C., and L.C. performed the statistical analysis and assisted with the interpretation of results. J.H., N.T., L.W., G.F., S.S., and A.J. provided expertise in cardiovascular biology and GWAS summary statistics. All authors discussed the results and commented on the paper.

## Competing interests

All authors declare no competing interests.

## Nonstandard Abbreviations and Acronyms

RT: respiratory diseases
CVD: cardiovascular diseases
SNP: single nucleotide polymorphisms
GWAS: genome-wide association studies
MR: Mendelian randomization
COPD: chronic obstructive pulmonary disease
IPF: idiopathic pulmonary fibrosis
OSA: sleep apnoea
AF: atrial fibrillation
CAD: coronary artery disease
VTE: venous thromboembolism
HF: heart failure
PAD: peripheral artery disease
MAGMA: multi-marker analysis of genomic annotation
eMAGMA: eQTL Multi-marker Analysis of GenoMic Annotation
SMR: Summary data-based Mendelian Randomization
GBMI: Global Biobanking Meta-Analysis Initiative
MAF: minor allele frequency
LDSC: Linkage disequilibrium score
rgs: genetic correlation
MHC: major histocompatibility complex
LDSC-SEG: LDSC applied to specificaly expressed genes
AIC: Akaike Information Criterion
LAVA: Local Analysis of [co]Variant Association
PLACO: pleiotropy analysis under composite null hypothesis
FUMA: functional mapping and annotation
LD: linkage disequilibrium
CADD: combined annotation dependent depletion
RegulomeDB: probability of regulatory functionality
PPH4: posterior probability of H4
TWAS: transcriptome-wide association study
eQTL: expression quantitative trait loci
FUSION: Functional Summary-based ImputatiON
BLUP: best linear unbiased prediction
BSLMM: modeling SNPs and effect sizes
HEIDI: heterogeneity of associated instrumentation
FDR: false discovery rate
BH: Benjamini-Hochberg
LHC-MR: Latent Heritable Confounder MR
InSIDE: instrument strength independent of direct effect
IVW: inverse variance weighting
MR-PRESSO: MR-pleiotropy residual sum and outlier
LCV: Latent Causal Variable
GCP: genetic causality proportion
TNFSF12: tumor necrosis factor ligand superfamily member 12
TWEAK: TNF-related weak inducer of apoptosis
HASMC: human airway smooth muscle cells

## Figure Legend

**Figure 1:**
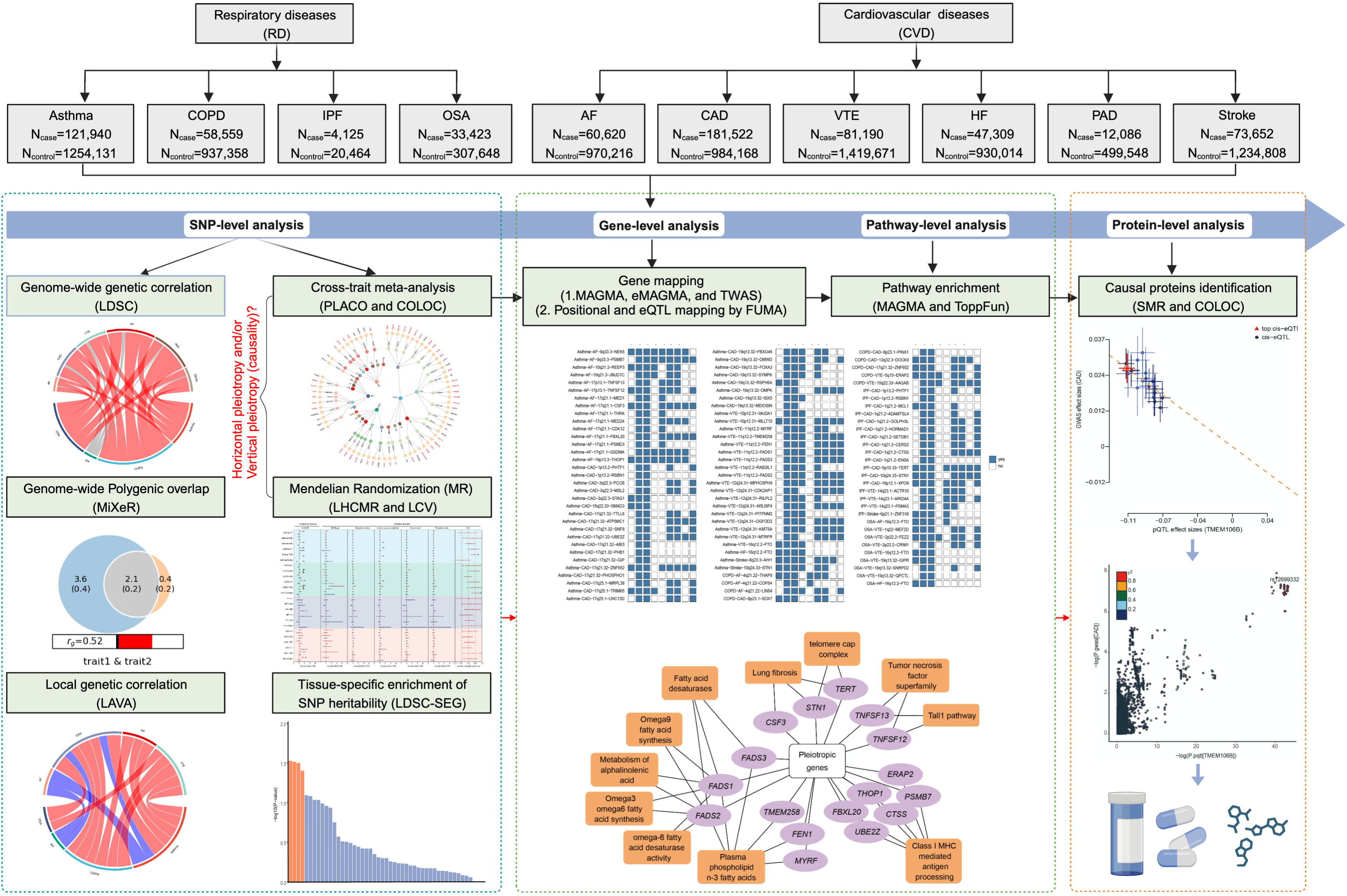
Overall study design of genome-wide cross-trait analysis. This figure illustrates the comprehensive analytical approach undertaken for 4 respiratory diseases and 6 cardiovascular diseases within this study. Initial GWAS findings were sourced from various repositories. Preceding the integration of respiratory and cardiovascular diseases in a multi-trait analysis, genetic characteristics such as SNP-based heritability, genome-wide or local genetic correlations, polygenic overlap were estimated individually. Analysis of SNPs and genomic loci across trait assessments uncovers novel pleiotropic loci for respiratory and cardiovascular diseases. To comprehensively characterize the genetic architecture of each respiratory and cardiovascular disease, multiple approaches (positional and eQTL mapping) were used to identify genes associated with pleiotropic loci and explore their biological pathways. Overlap within circulating proteins is then assessed through proteome-wide Mendelian randomization and colocalization analysis, which provides preclinical indicators for drug development strategies. Finally, Mendelian randomization analysis was used to evaluate potential causal relationships between respiratory and cardiovascular diseases.

**Figure 2:**
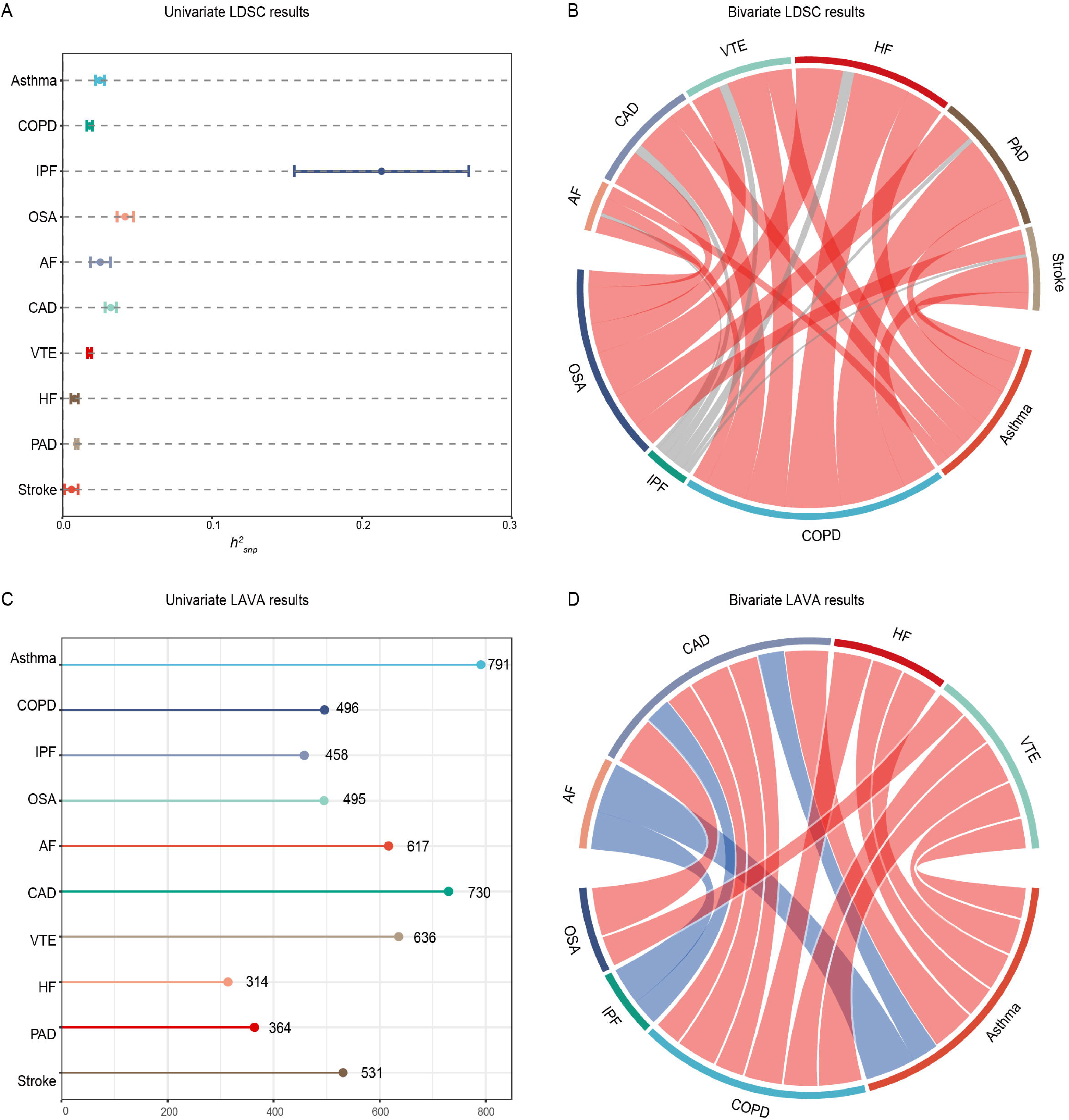
SNP-based heritability and polygenic overlap of the four respiratory diseases and six cardiovascular diseases. (A) Error-bar plot of the SNP-based heritability (h2SNP) point estimates for the four respiratory diseases and six cardiovascular diseases, computed using univariate LDSC. (B) Network visualization of the Bonferroni-corrected significant global r_g_s among the respiratory and cardiovascular diseases, computed using bivariate LDSC. Connections represent significant r_g_s, with correlation value along connections, thicker lines denoting stronger correlations, and red indicates correlation between trait pairs passing Bonferroni corrected thresholds. The size of the nodes is weighed by the sample size and h^2^_SNP_ of the given respiratory and cardiovascular disease (size = h^2^_SNP_ × sqrt(N)); (C) Lollipop plot of the number of sufficient local genetic signals for the four respiratory diseases and six cardiovascular diseases, computed using univariate LAVA; (D) Network visualization of the Bonferroni-corrected significant local r_g_s between pairs of respiratory and cardiovascular diseases, computed using bivariate LAVA. Connections represent how many significant local r_g_s were identified between pairs of respiratory and cardiovascular diseases, with thicker connections denoting higher numbers. Red represents positive local r_g_s, while blue represents negative local rgs. The orientation and the size of the nodes were set to mirror that of the network visualization of global r_g_s.

**Figure 3:**
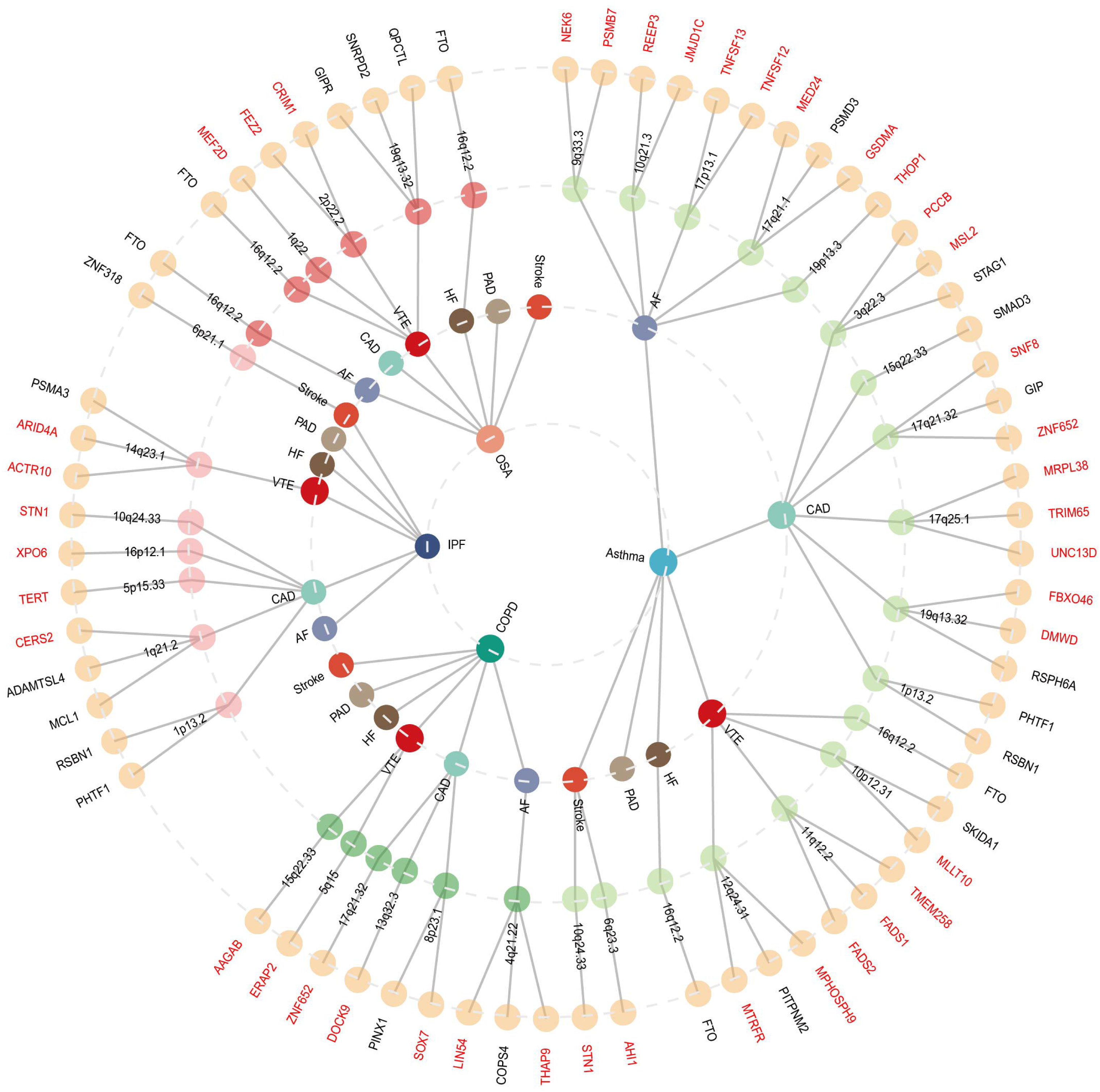
The overall landscape of the pleiotropic associations across four Respiratory Diseases and six Cardiovascular Diseases. A circular dendrogram included four respiratory diseases (inner circle, including Asthma, COPD, IPF, and OSA) and six cardiovascular diseases (AF, CAD, VTE, HF, PAD, and Stroke), resulting in 24 trait pairs. A total of 97 pleiotropic loci were identified across 14 trait pairs (third circle; no pleiotropic loci were identified for asthma-PAD, COPD-HF, COPD-PAD, COPD-Stroke, IPF-AF, IPF-HF, IPF-PAD, OSA-CAD, OSA-PAD, and OSA-Stroke). A total of 97 significant pleiotropic genes (41 unique) were further identified by multimarker analysis of GenoMic annotation (MAGMA).Among the 97 risk genes identified by MAGMA, 59 genes were verified by the eMAGMA method based on eqtl and are marked in red. For the trait pairs with more than 3 pleiotropic genes, we only showed the top 3 pleiotropic genes according to the prioritization of candidate pleiotropic genes (fourth circle).

**Figure 4:**
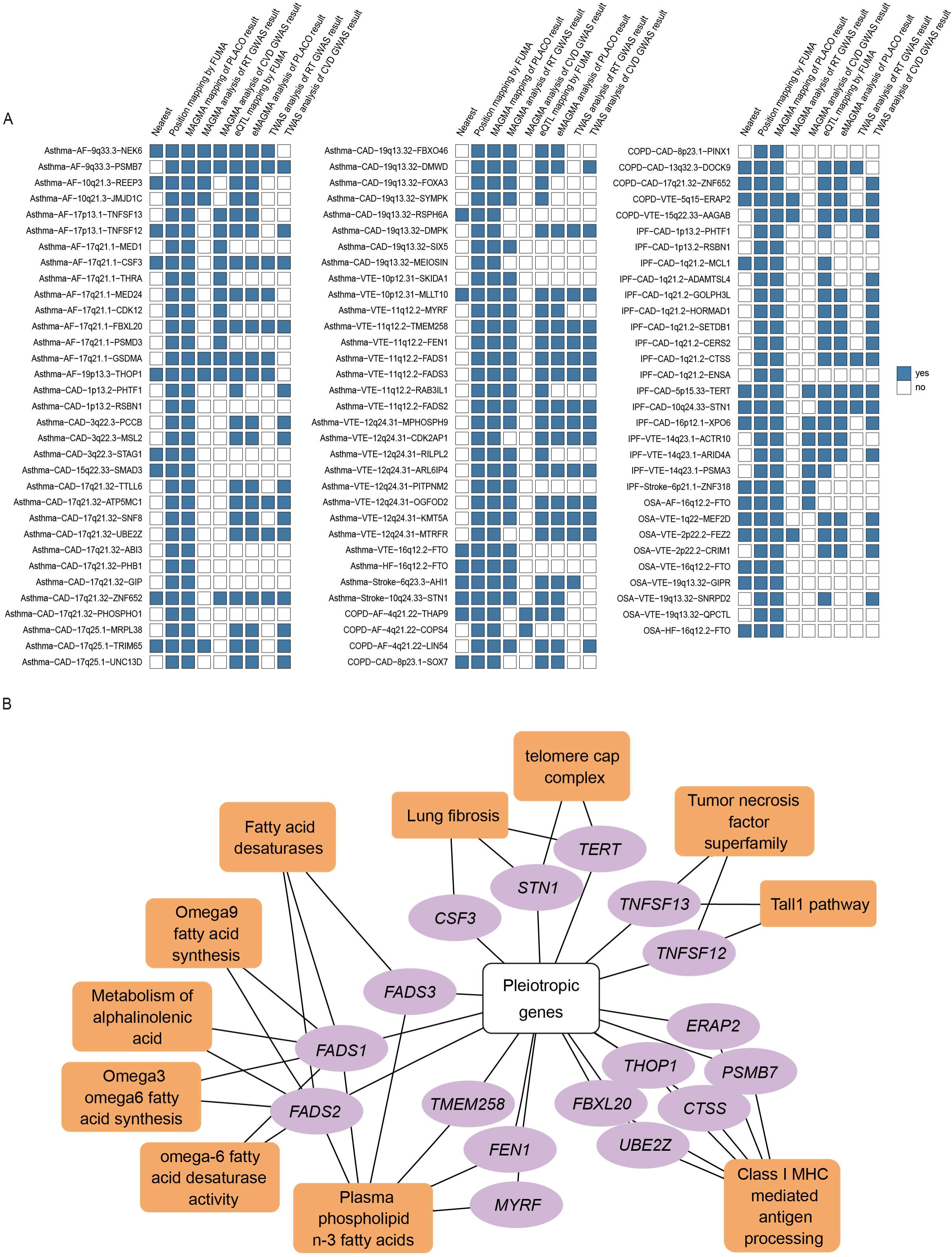
Overall landscape of pleiotropic genes identified by both MAGMA and eMAGMA. (A) The heatmap represents the location of 59 pleiotropic genes jointly identified by MAGMA and eMAGMA in PLACO, MAGMA and TWAS. Blue indicates that the gene was mapped in the corresponding method. (B-C) Bar chart for GO BP and Reactome enrichment analysis.

**Figure 5:**
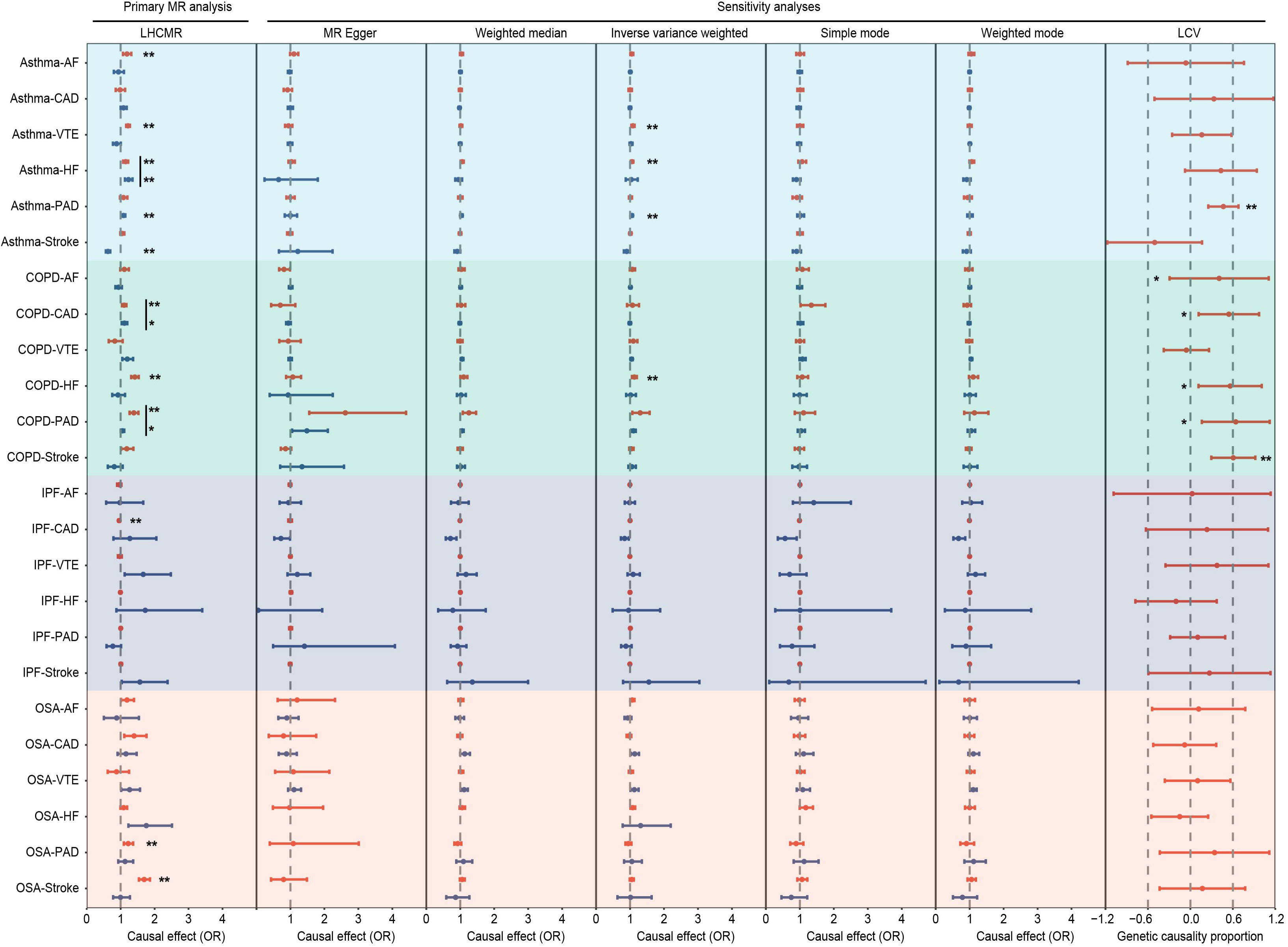
Mendelian randomization analysis of causal relationships between RTs and CVDs. LHC-MR plots for the association between 4 respiratory diseases and 6 cardiovascular diseases. The forward (RT→CVD) and reverse (CVD→RT) causal estimates are shown in two different colors as dots (red and blue). Sensitivity analysis including LCV further supplemented and verified the LHCMR method. * = significant effect after Boneferroni correction (i.e., *P*-value < 0.05); ** = significant effect after Boneferroni correction (i.e., *P*-value < 0.05/24).

**Figure 6:**
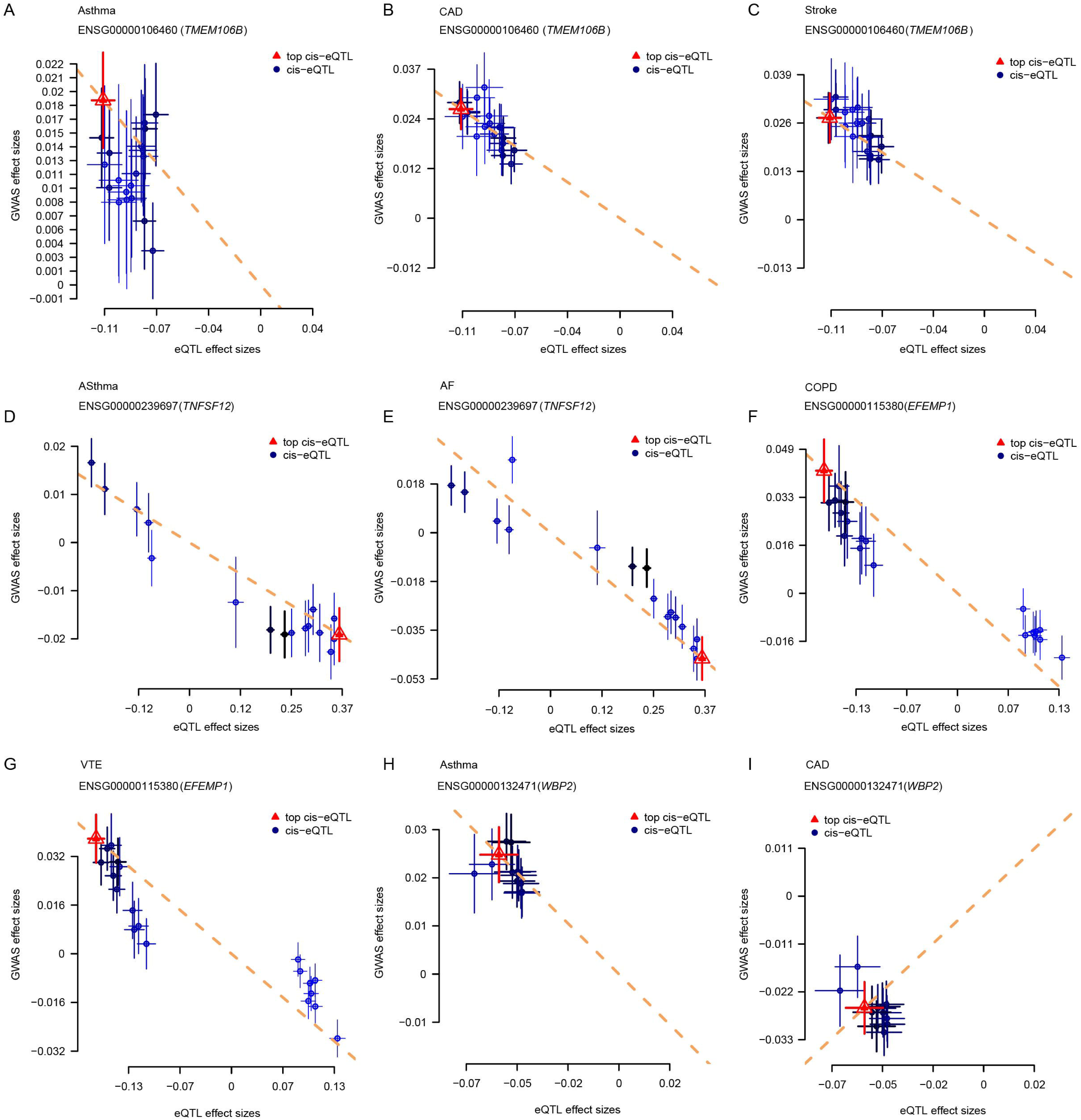
Three proteins that have an impact on both respiratory and cardiovascular diseases. Scatter plots showing that higher TMEM106B protein levels in the blood tend to have an increased risk of asthma, CAD and Stroke. Higher levels of TNFSF12 protein in the blood are associated with an increased risk of asthma and AF. Higher WBP2 protein in the blood increases the risk of asthma but reduces the risk of CAD. Each point represents a SNP, the x value of a SNP is its β effect size on a protein and the horizontal error bar represents the SE around the Beta. The y value of the SNP is its β effect size on disease risk and the vertical error bar represents the SE around its β. The dashed line represents the SMR estimate (a line with the intercept of 0 and the slope of β from the SMR test).

## Notes

### Competing Interest Statement

The authors have declared no competing interest.

