## Supplementary file for "Heart-lung connections: Phenotypic and genetic insights from a large-scale genome-wide cross-trait analysis"

**Supplementary Figures**

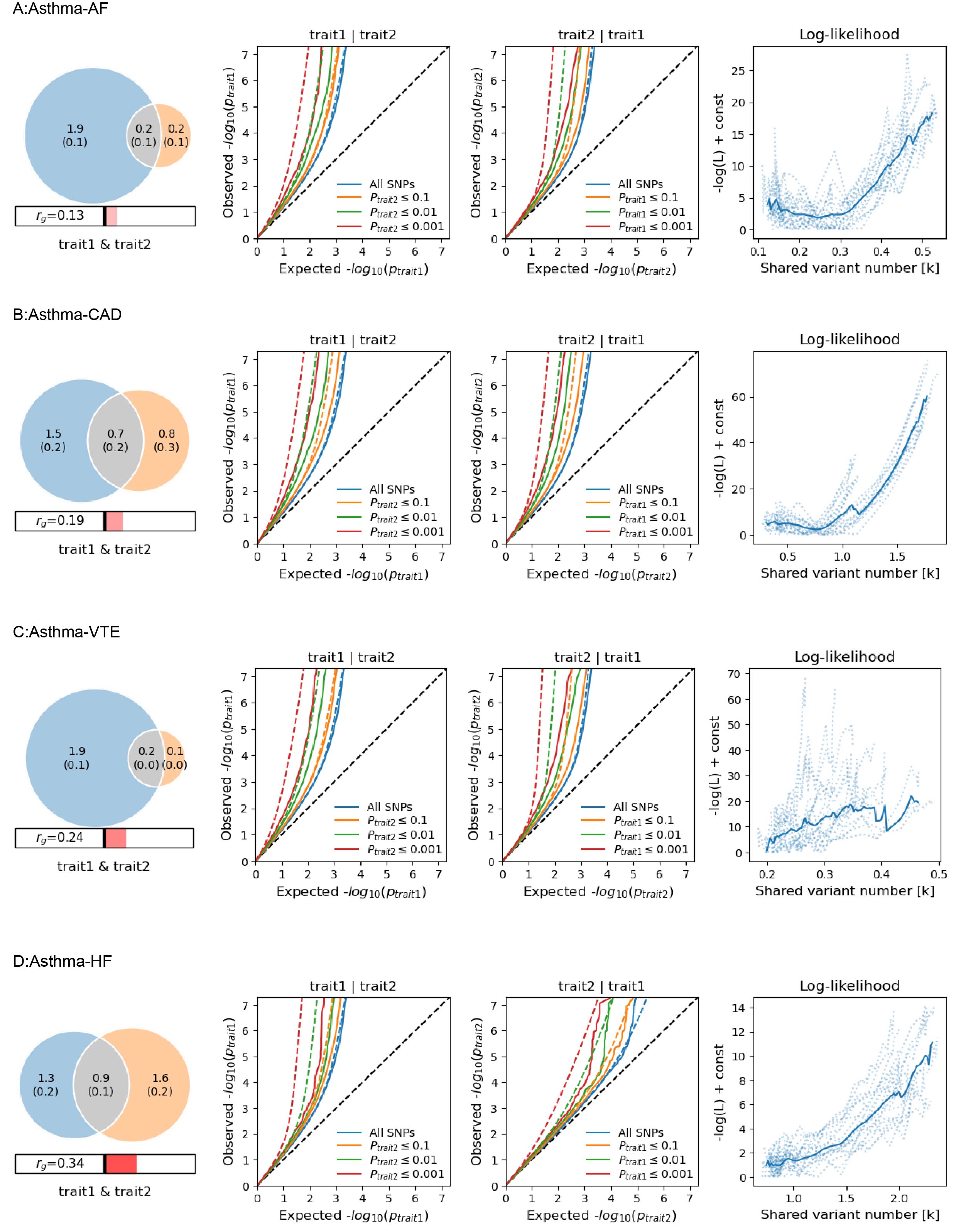

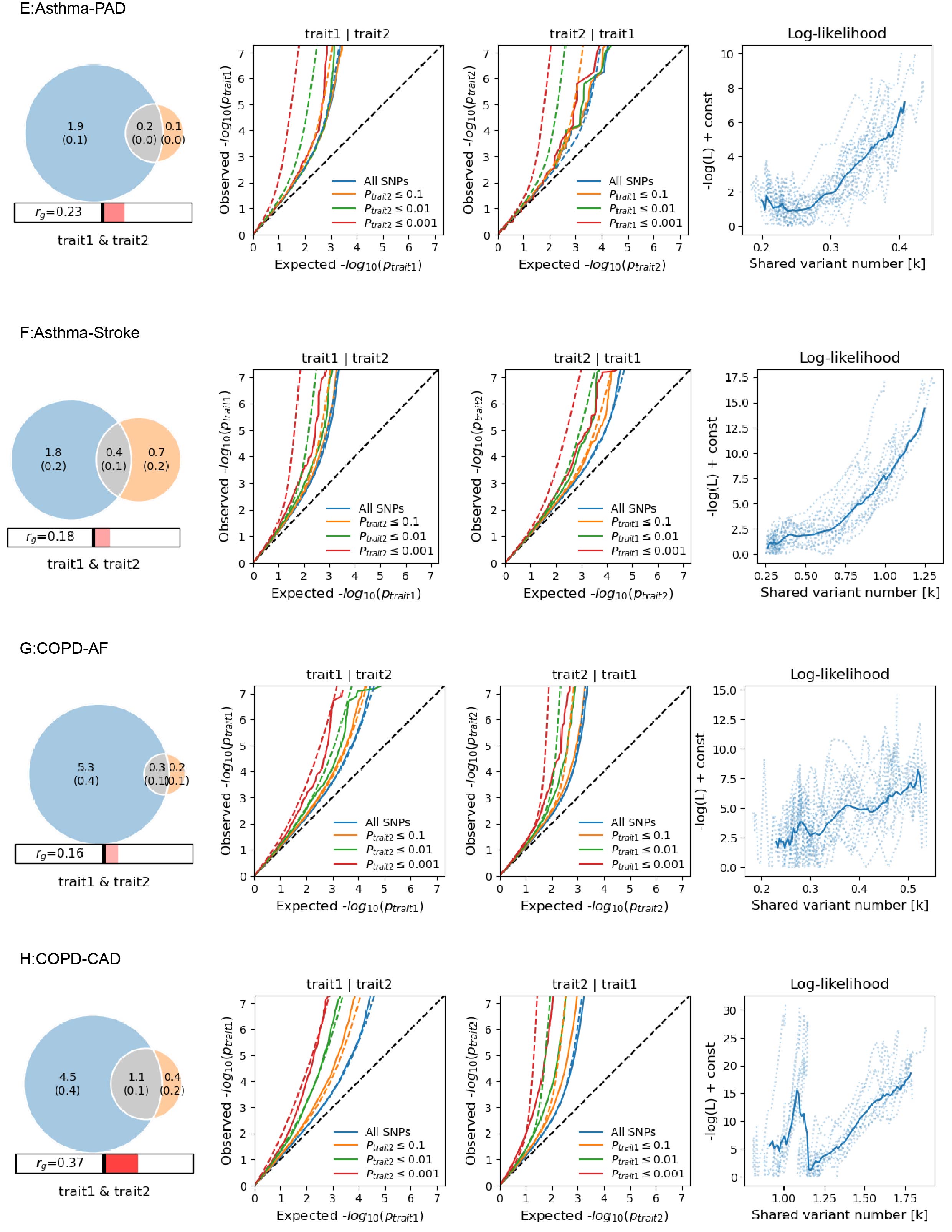

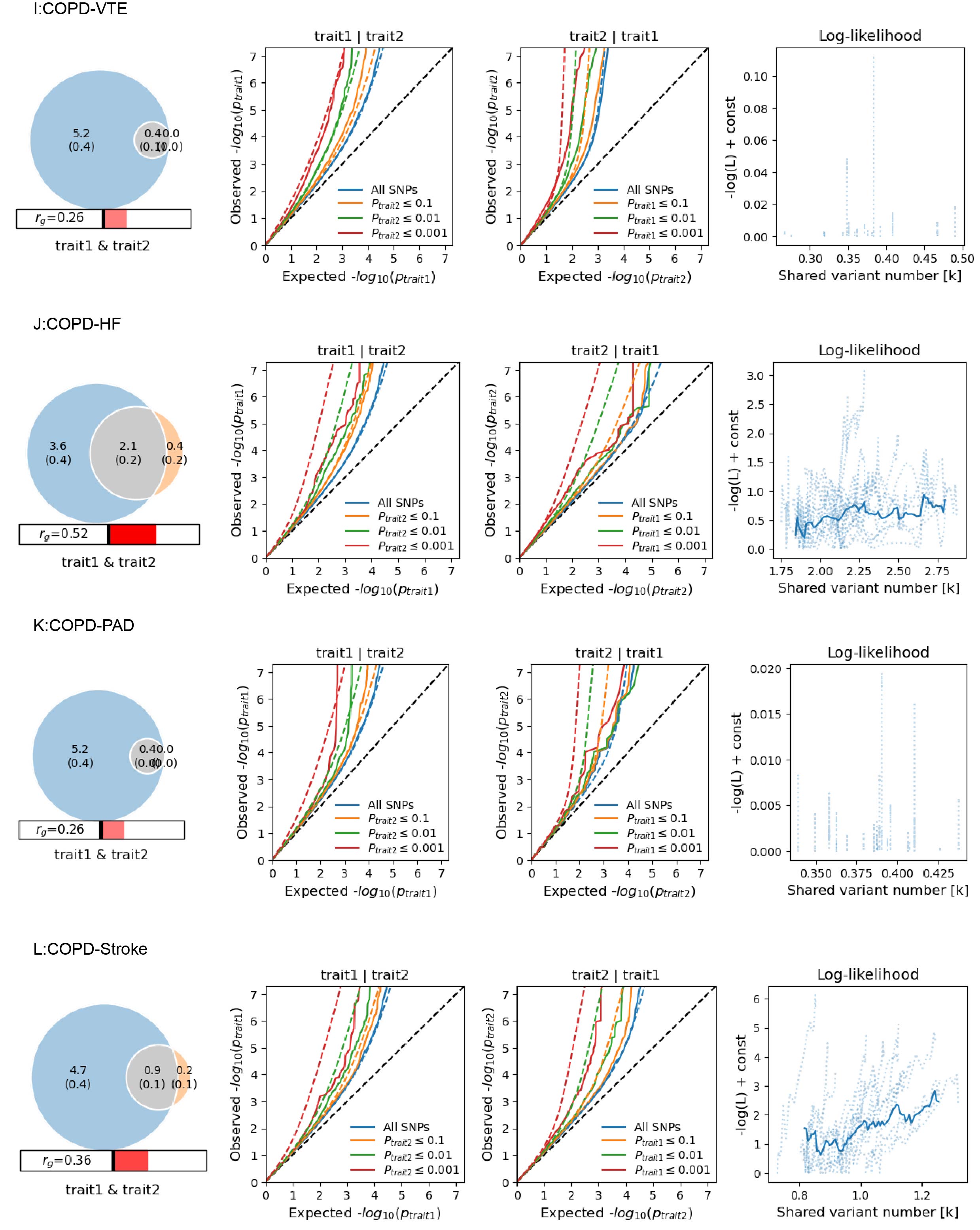

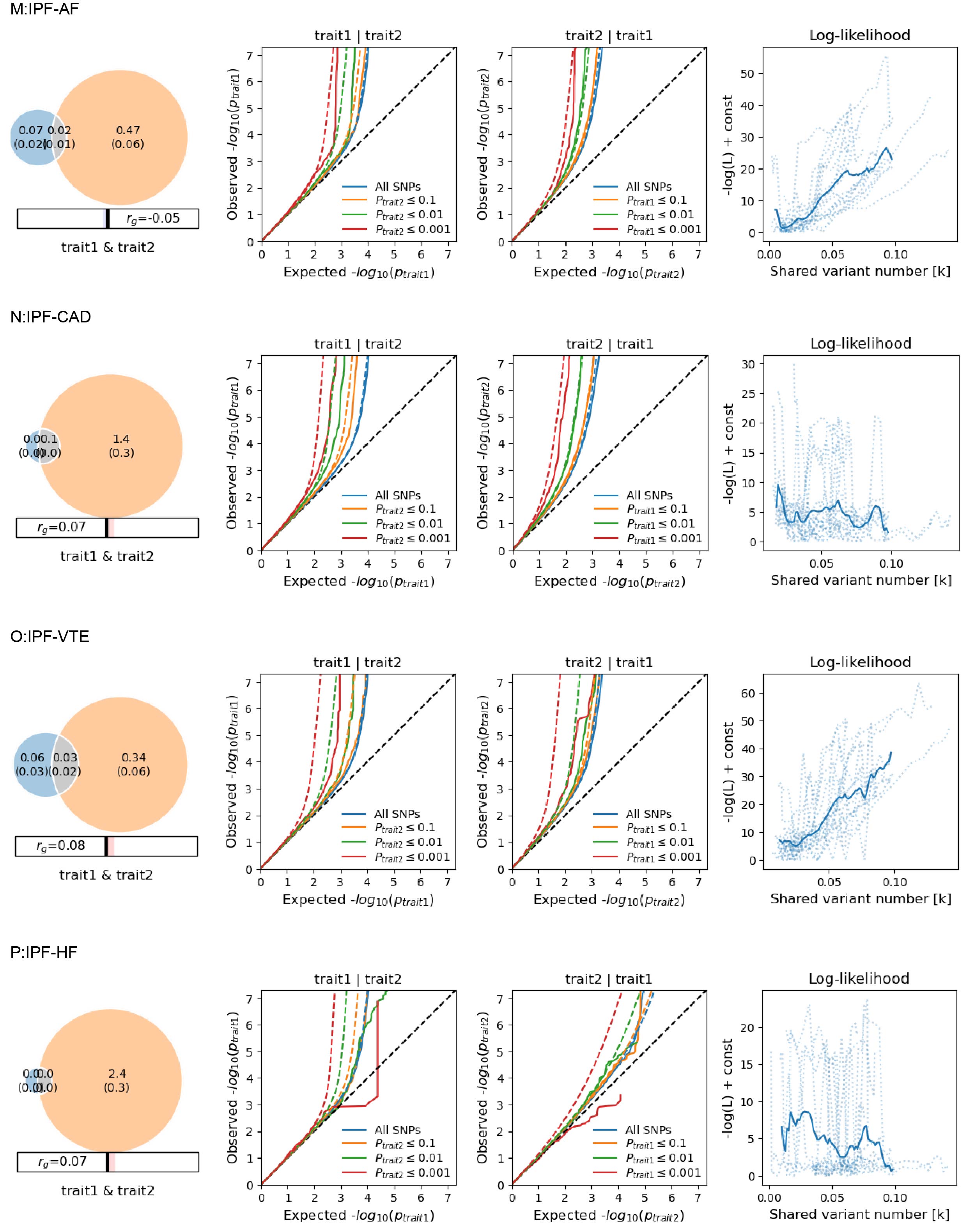

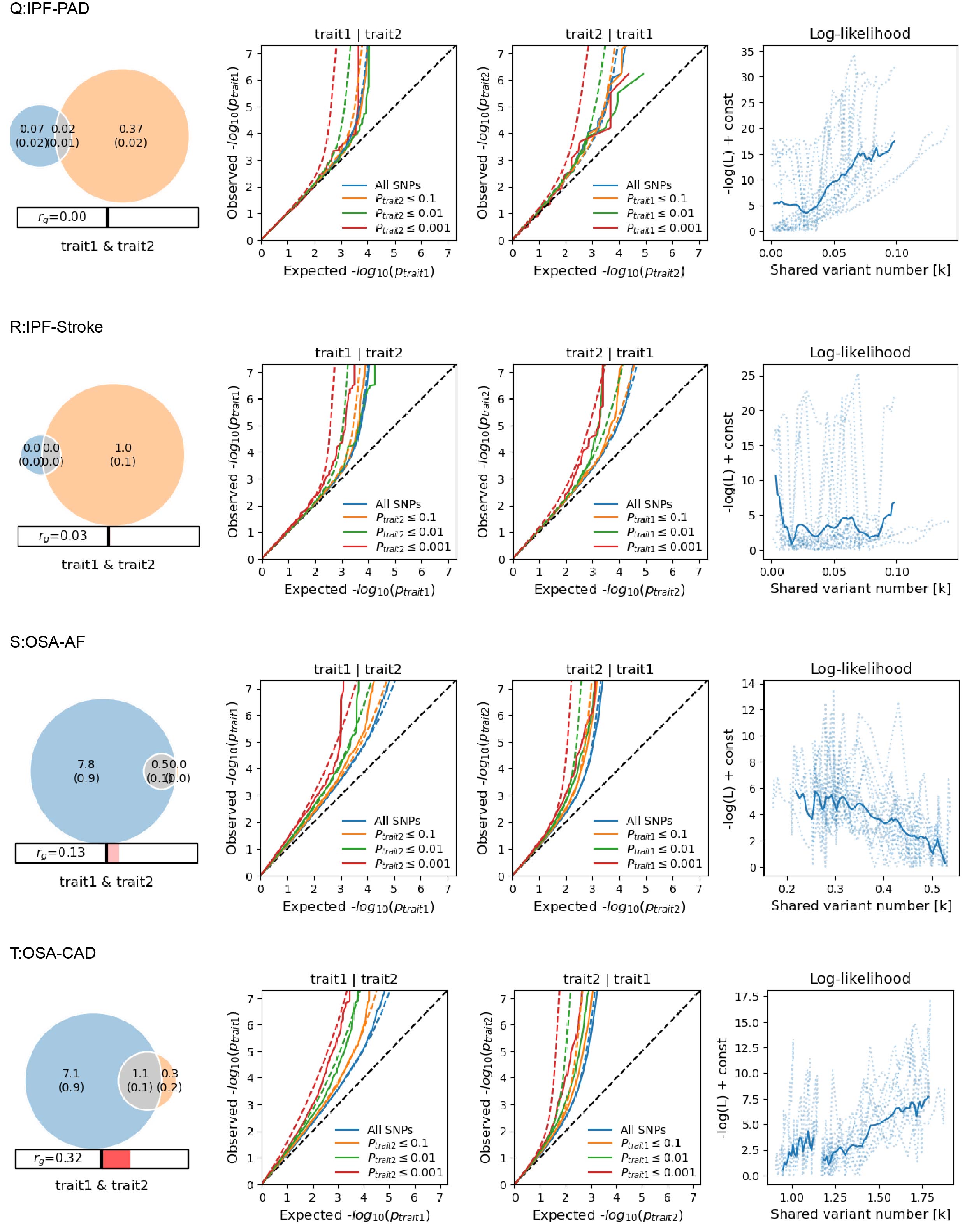

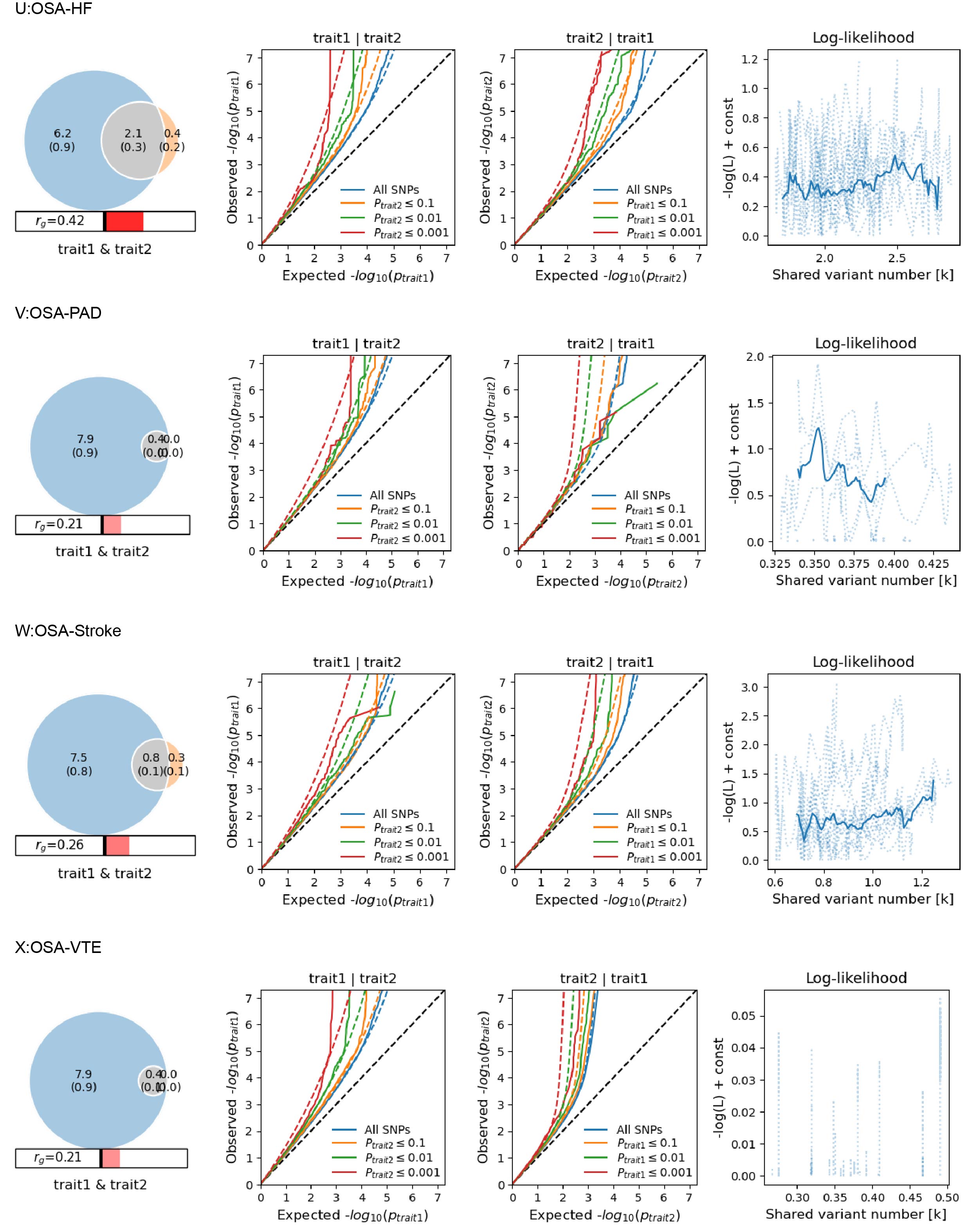

**Supplementary Fig. 1. Supplemental MiXeR figures for each of the 24 trait pairs of respiratory diseases and cardiovascular diseases.**

On the left, Venn diagrams depict the unique (blue and orange) and shared (grey) causal variants associated with pairs of respiratory diseases and cardiovascular diseases. Polygenic overlap is represented in gray. The numbers indicate the estimated quantity of causal variants (in thousands) per component, explaining 90% of SNP heritability in each pair of respiratory diseases and cardiovascular diseases, with the standard error in parentheses. The size of the circles reflects the degree of polygenicity for each trait. The estimated genetic correlation for pairs of respiratory diseases and cardiovascular diseases is also shown below the corresponding Venn diagram, with an accompanying directional scale (blue shades for negative values and red shades for positive values).In the middle, conditional QQ plots of observed versus expected -log10 p-values in the primary trait as a function of the significance of the association with the secondary trait at the level of all SNPs (blue lines), p ≤ 0.1 (orange lines), p ≤ 0.01 (green lines), and p ≤ 0.001 (red lines). Dotted lines indicate model predictions for each stratum. The black dotted line is the expected Q-Q plot under the null hypothesis (no SNPs associated with the phenotype). Points on the Q-Q plot are weighted according to LD structure using n=64 iterations of random pruning at an LD threshold of r2 = 0.1. On the right, log-likelihood curves highlight the goodness of model fit, by plotting the negative log-likelihood function (lower values correspond to better model fit) against the π12 parameter (number of influencing variants shared between two traits). The remaining parameters of the model were constrained to their fitted values. The π12 range on the log-likelihood plots goes from the smallest possible value π12 = rg*sqrt (π1u, π2u) that is still compatible with the estimated genetic correlation, up to the largest possible value π12 = min(π1u, π2u) that corresponds to the minimum total polygenicity among the two traits. The minimum point indicates the best-fitting model estimate of the number of influencing variants shared between two trait

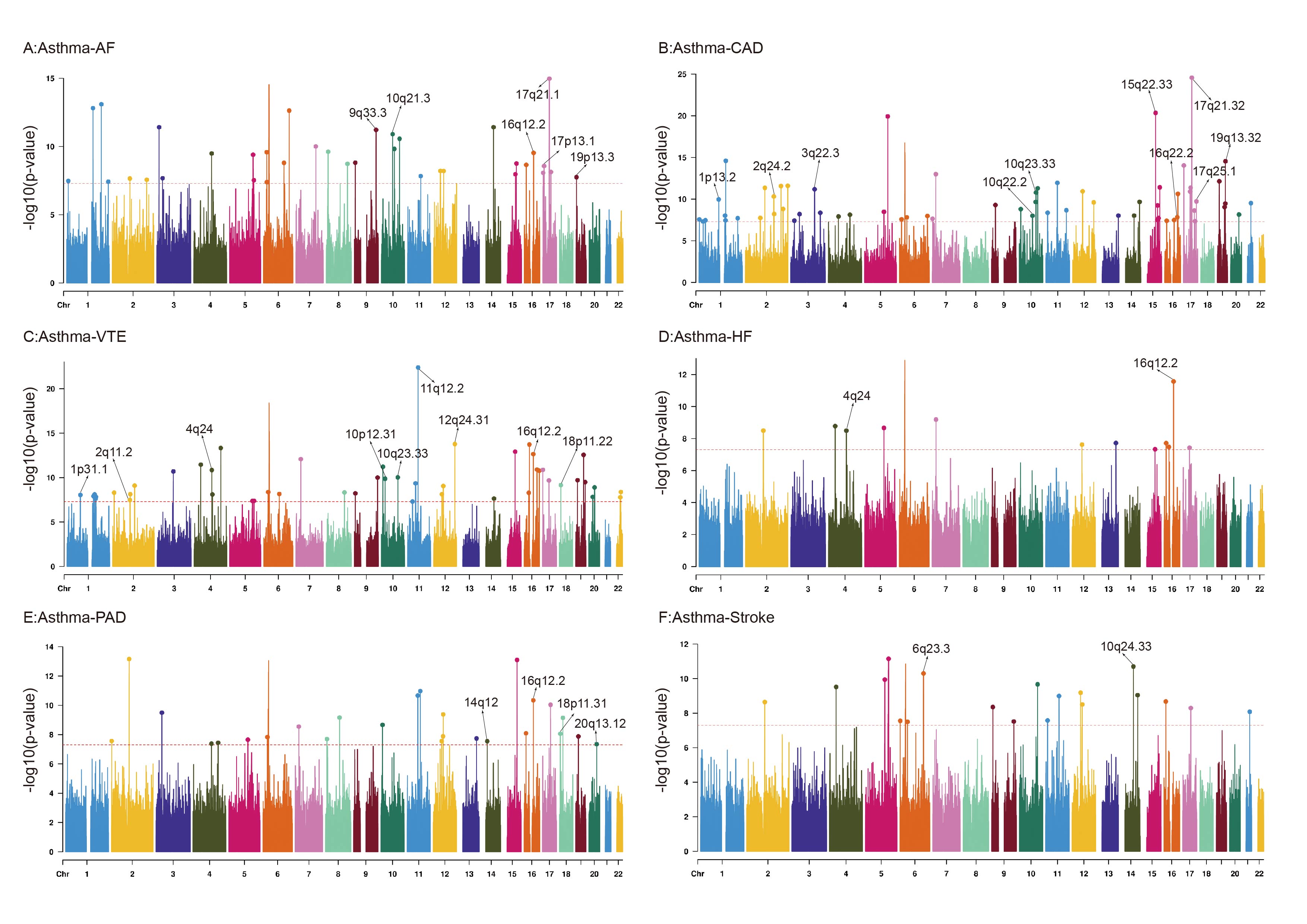

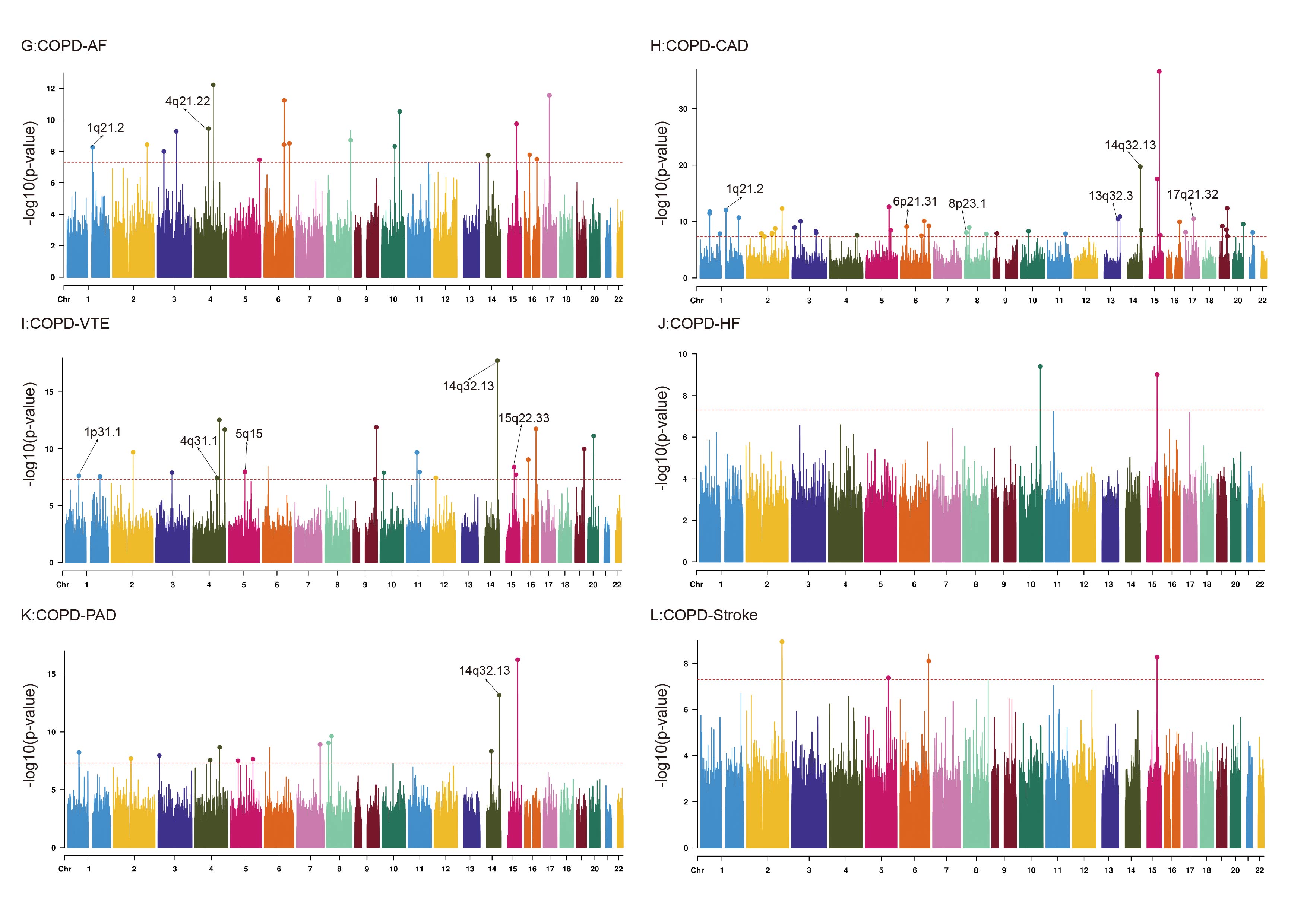

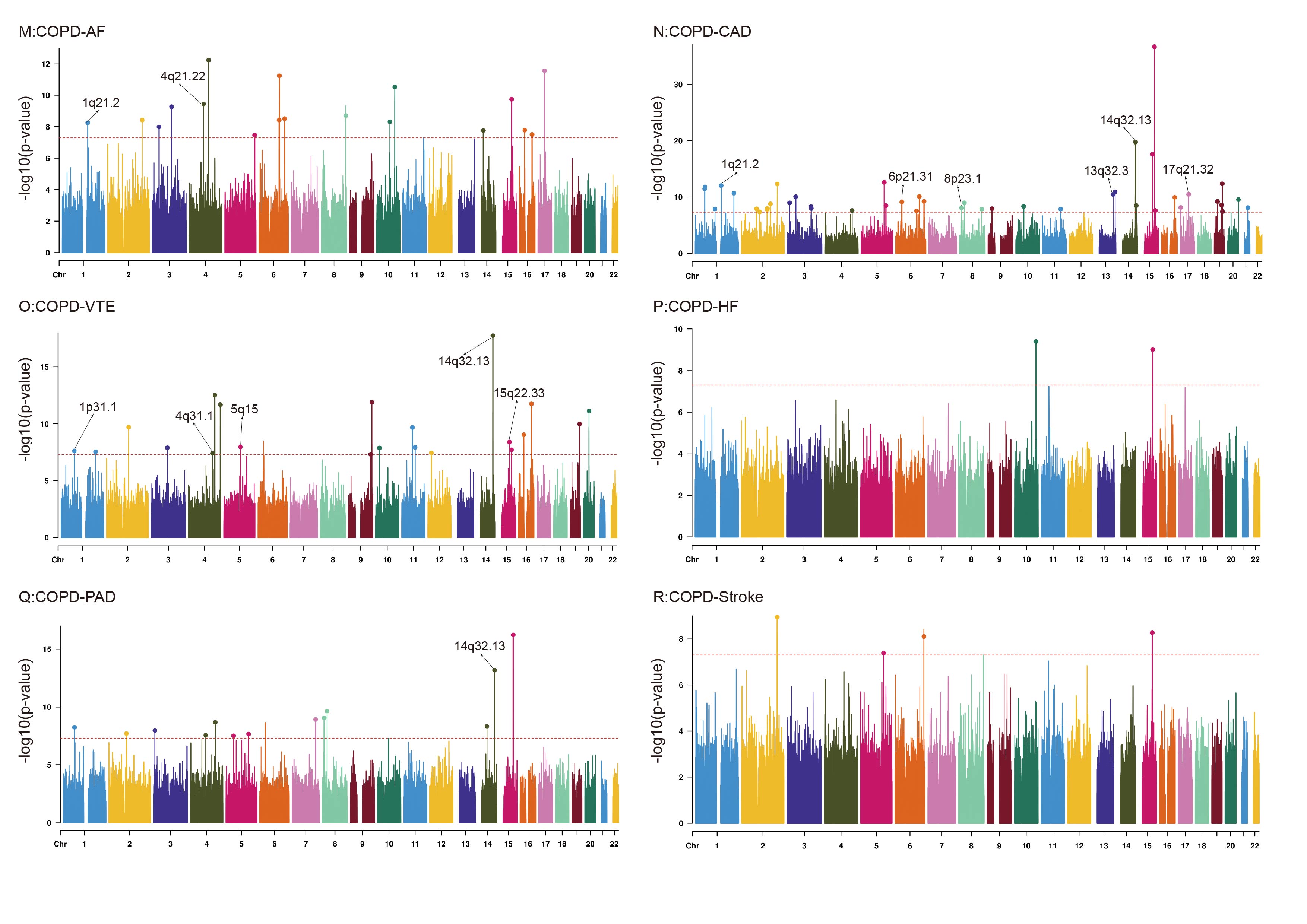

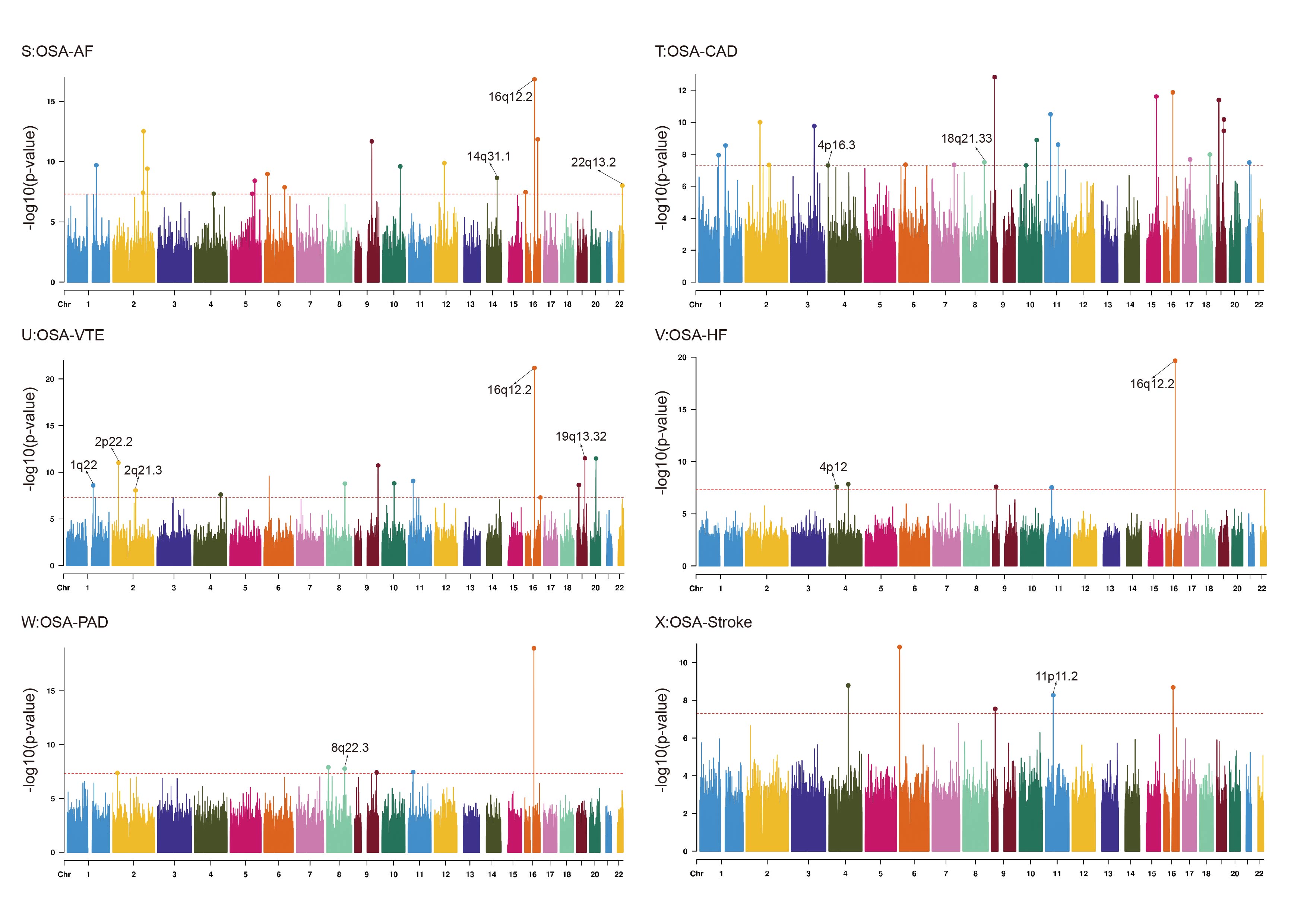

**Supplementary Fig. 2. Manhattan plots for the PLACO results of each of the 24 trait pairs.**

The X-axis is the chromosomal position, and the Y-axis is negative log10 transformed *P*-values for each SNP. The cytoband annotations for the newly identified genomic loci are in gray. The dotted lines in red indicate the genome-wide significant *P*-value of −log10 (5×10^−8^). Colorful dots indicate an independent genome-wide significant association with the smallest *P*-value (Top lead SNP). Only SNPs shared across all summary statistics were included.

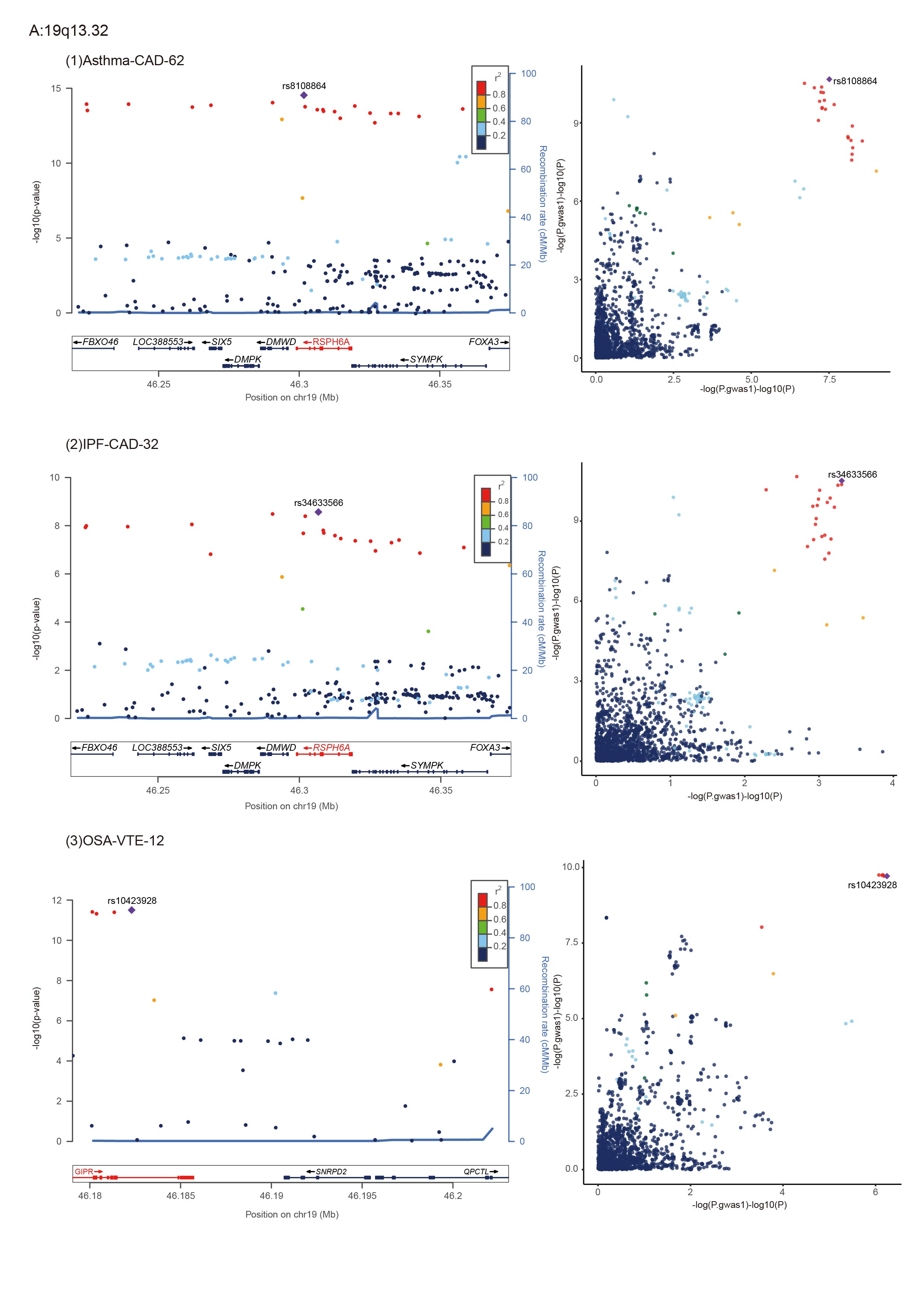

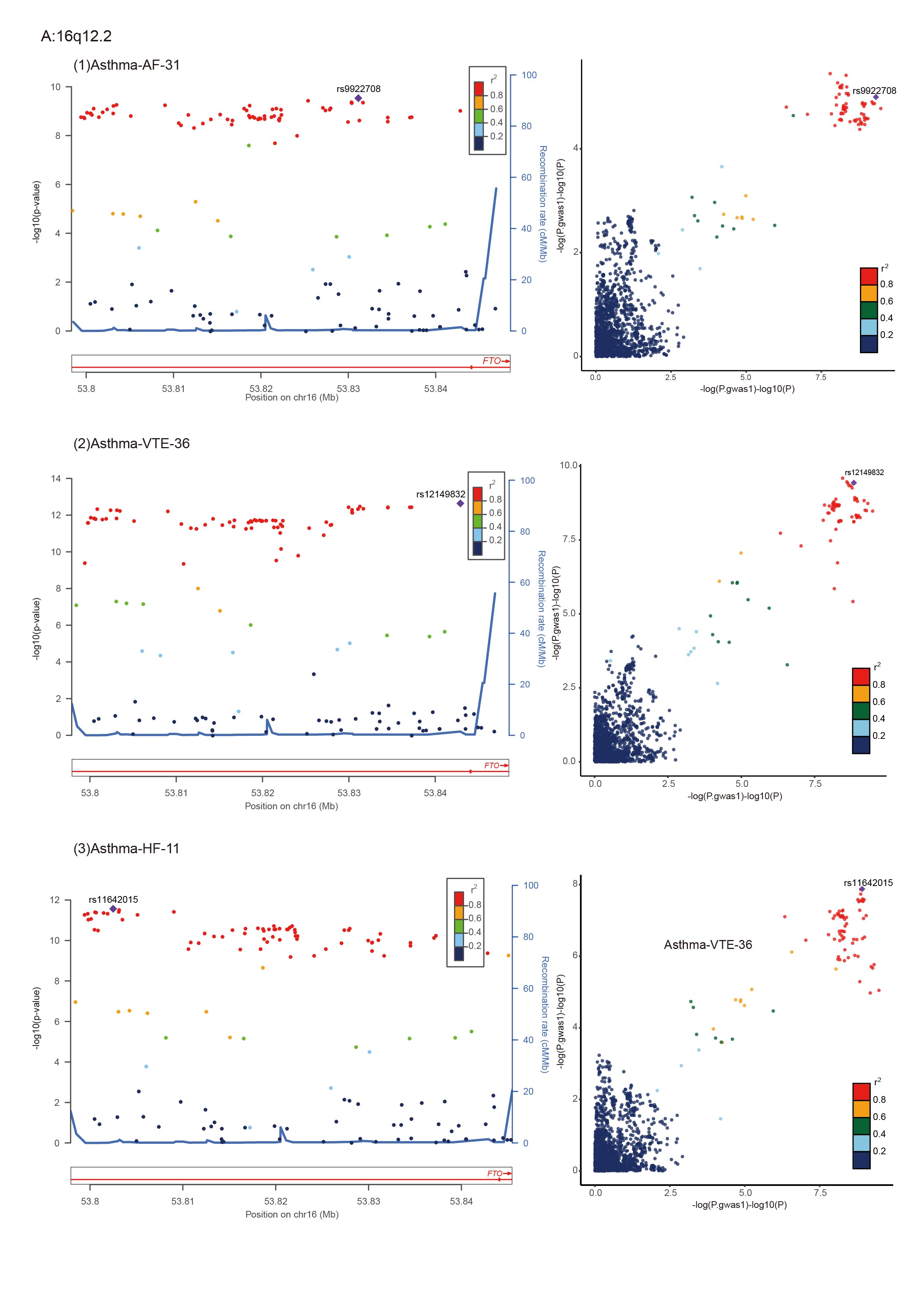

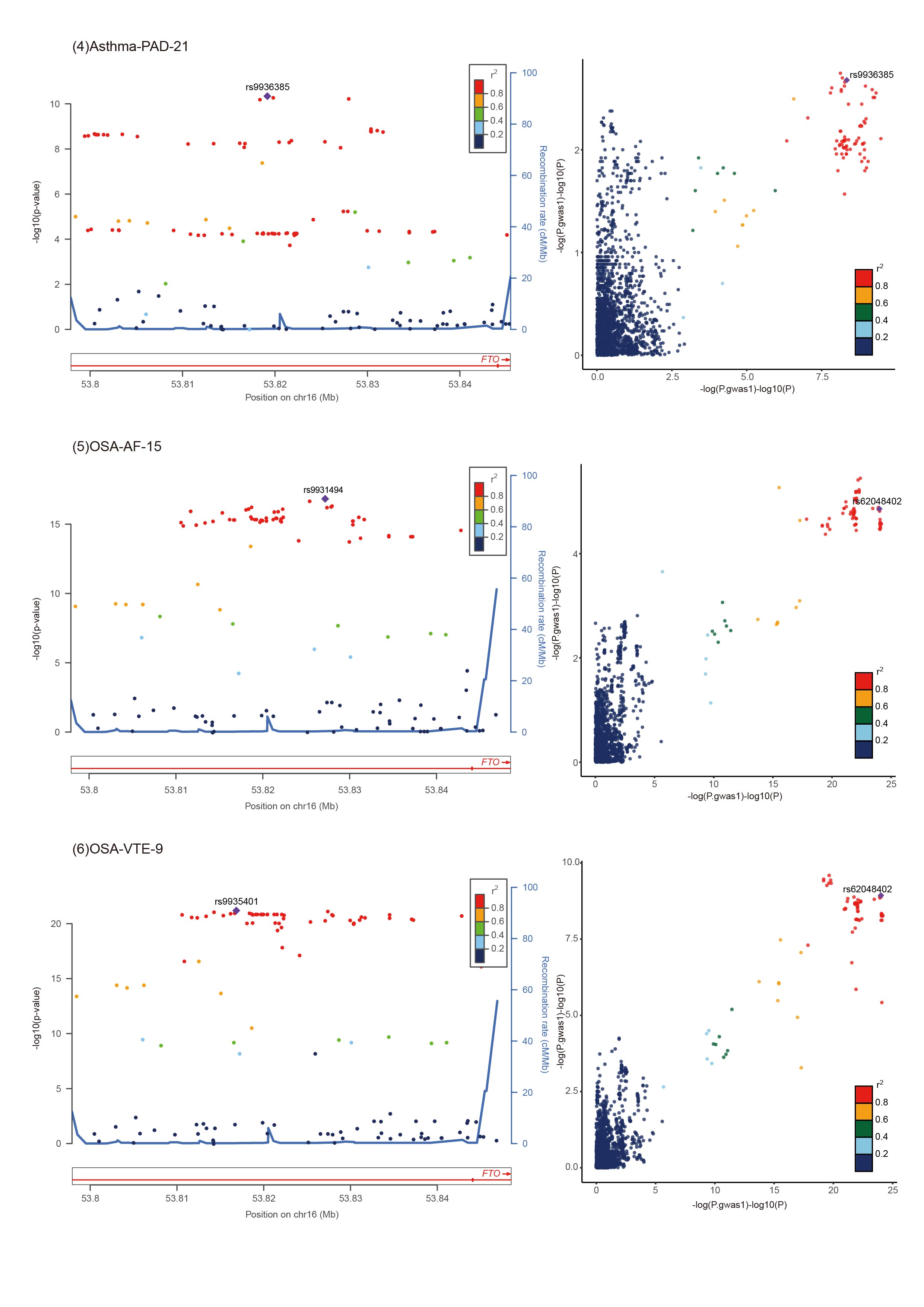

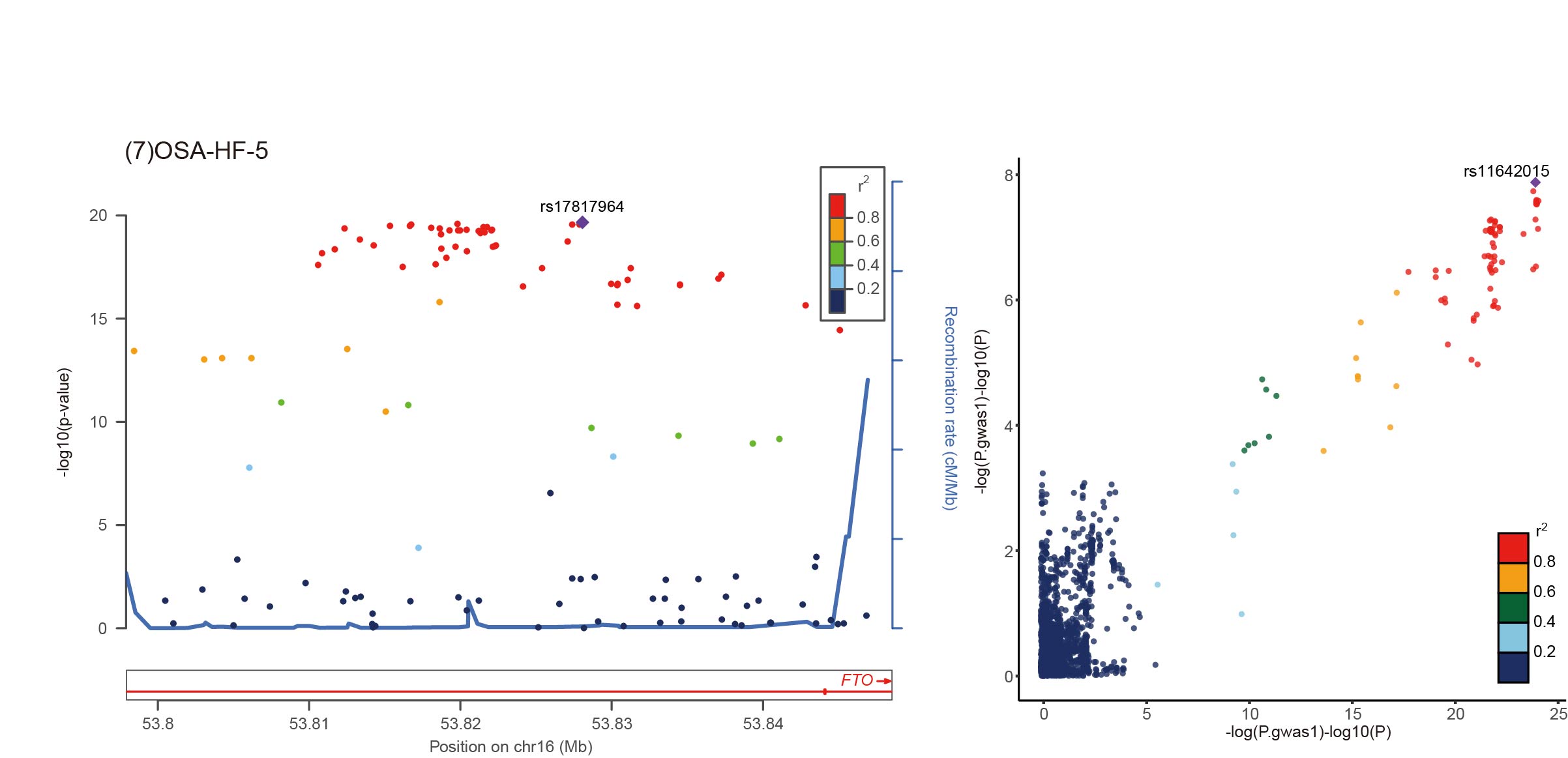

**Supplementary Fig. 3. LocusZoom and LocusCompare Plots of 10 Significantly Colocalized Loci in 2 Significant Regions**

For the significant colocalized locus (PP.H4 > 0.7) in regions 19q13.32 and 16q12.2 identified for corresponding trait pair, the left panel depicts the PLACO results using LocusZoom plot, and the right panel compares two single-trait GWAS statistics of the corresponding trait pair for each variant using LocusCompare plot. For the LocusZoom plot, the x-axis shows the genomic position for each variant, and the y-axis shows negative log10 P values from PLCAO results. The top variant with the smallest PPLACO in each locus is indicated in purple diamond. The color of each variant represents its LD relationship with the top variant.The linkage disequilibrium (LD) relationship between the lead SNP and the surrounding SNPs is indicated by the r2 legend.For the LocusCompare plot,each dot represents a variant, the x-axis shows the −log10 PGWAS from the corresponding GWAS of respiratory diseases, and the y-axis shows −log10 PGWAS from the corresponding GWAS cardiovascular diseases.

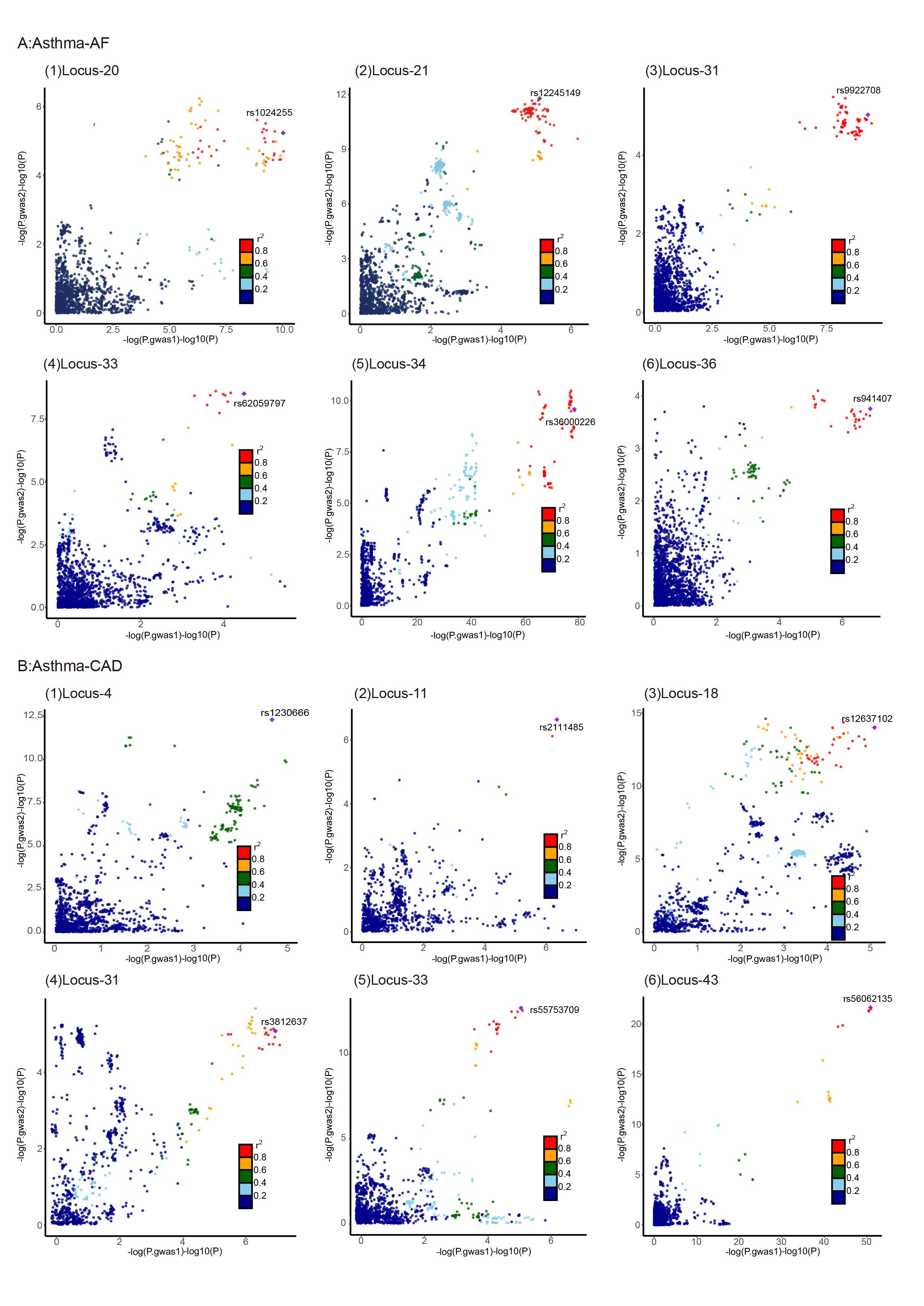

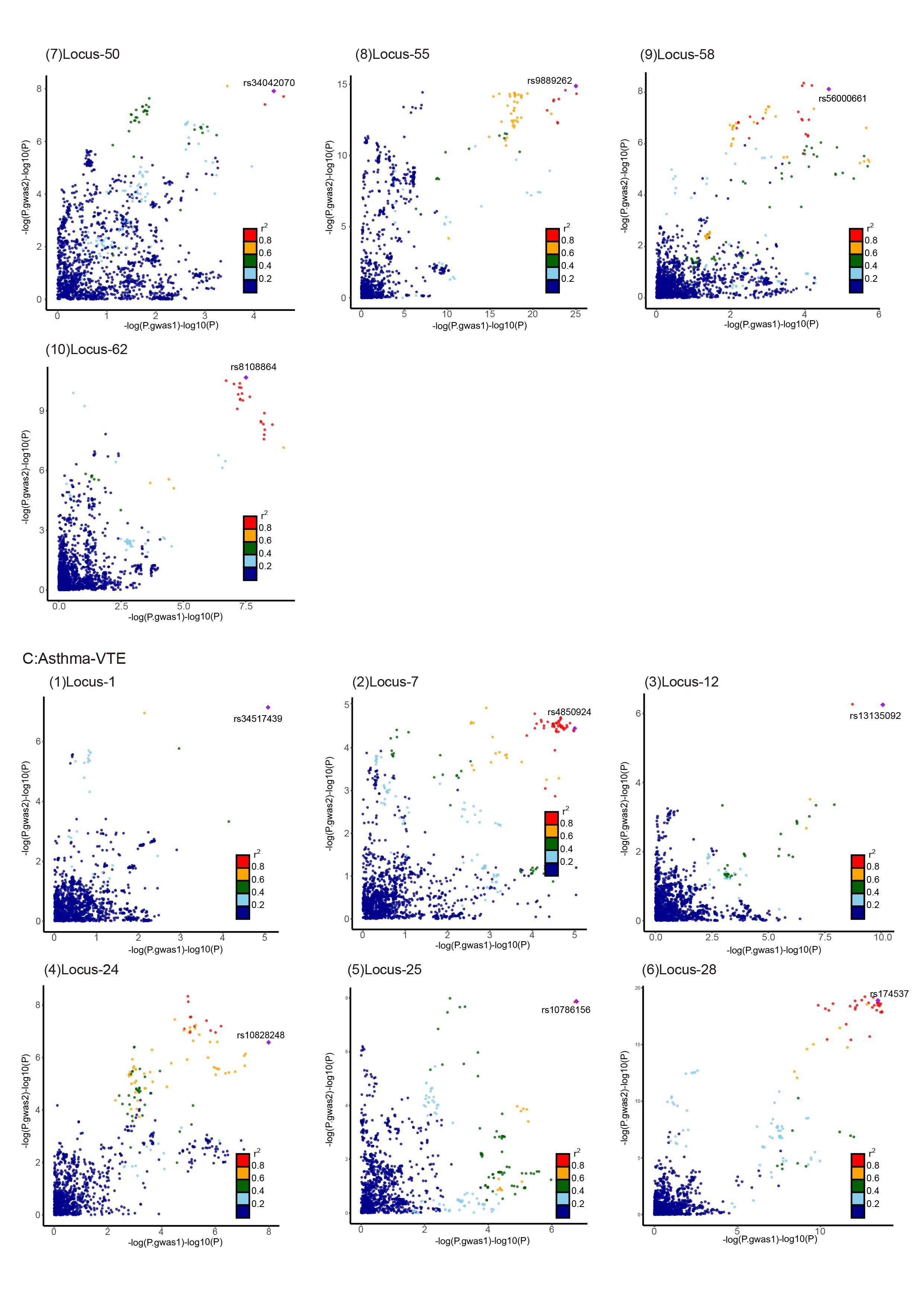

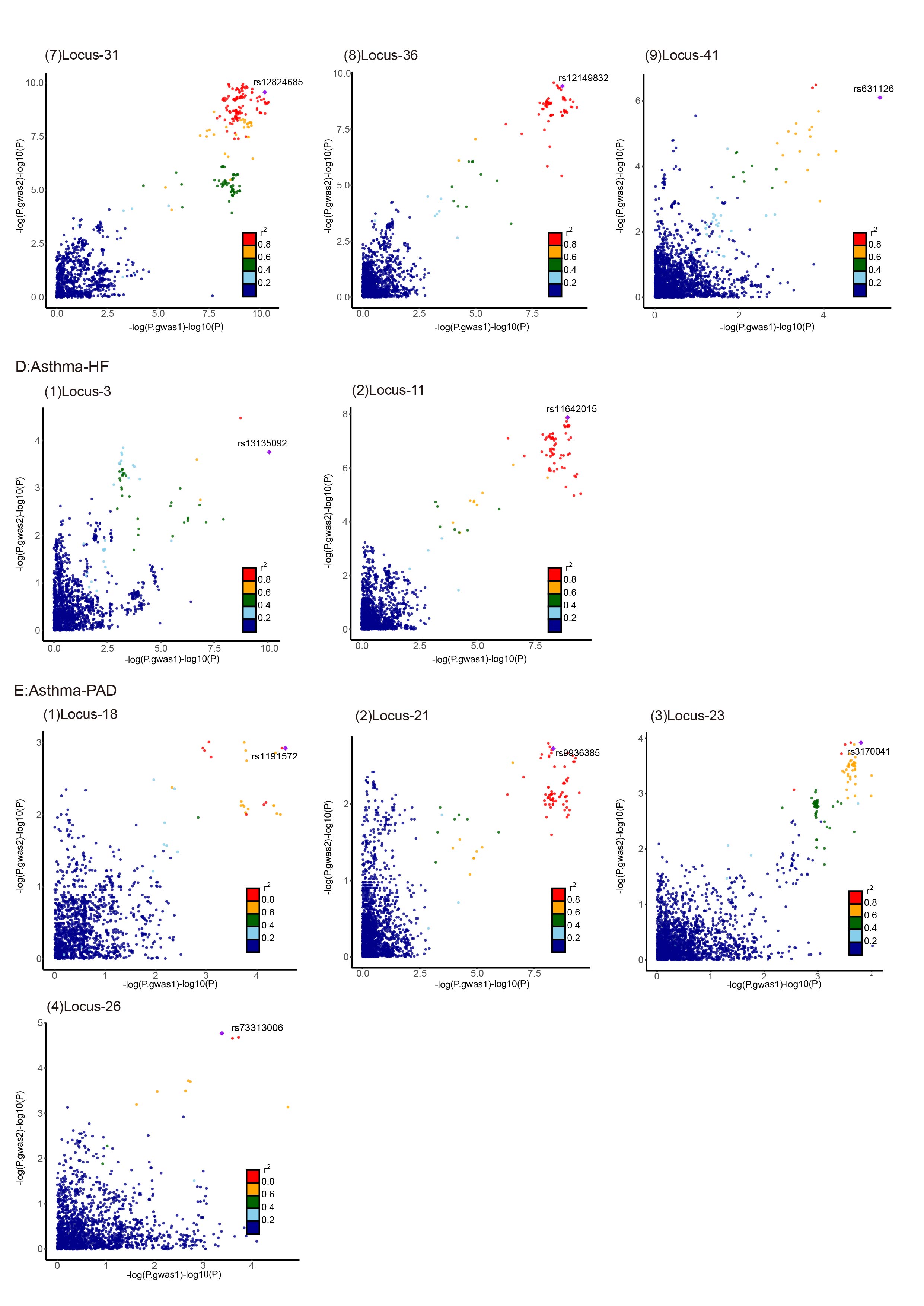

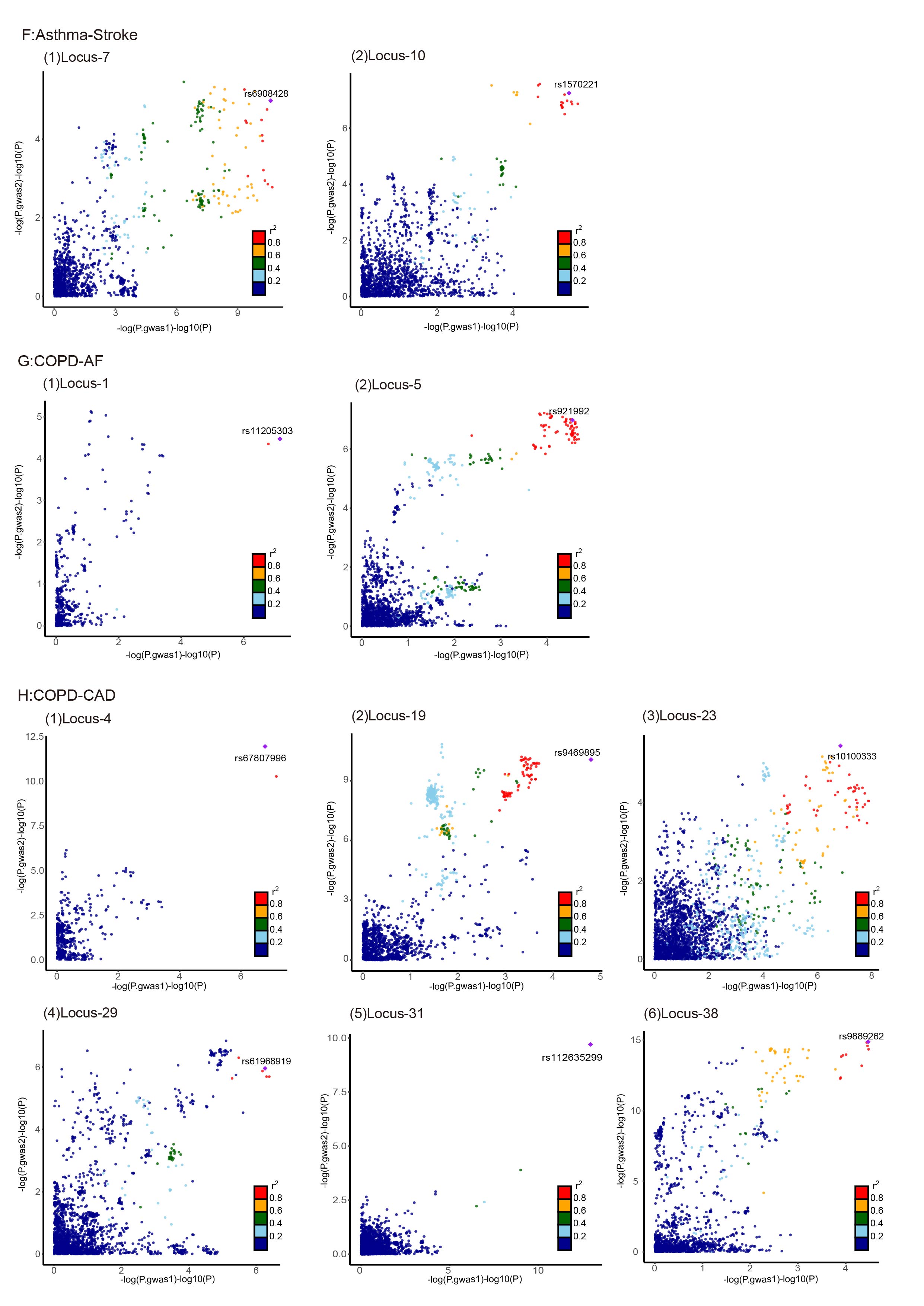

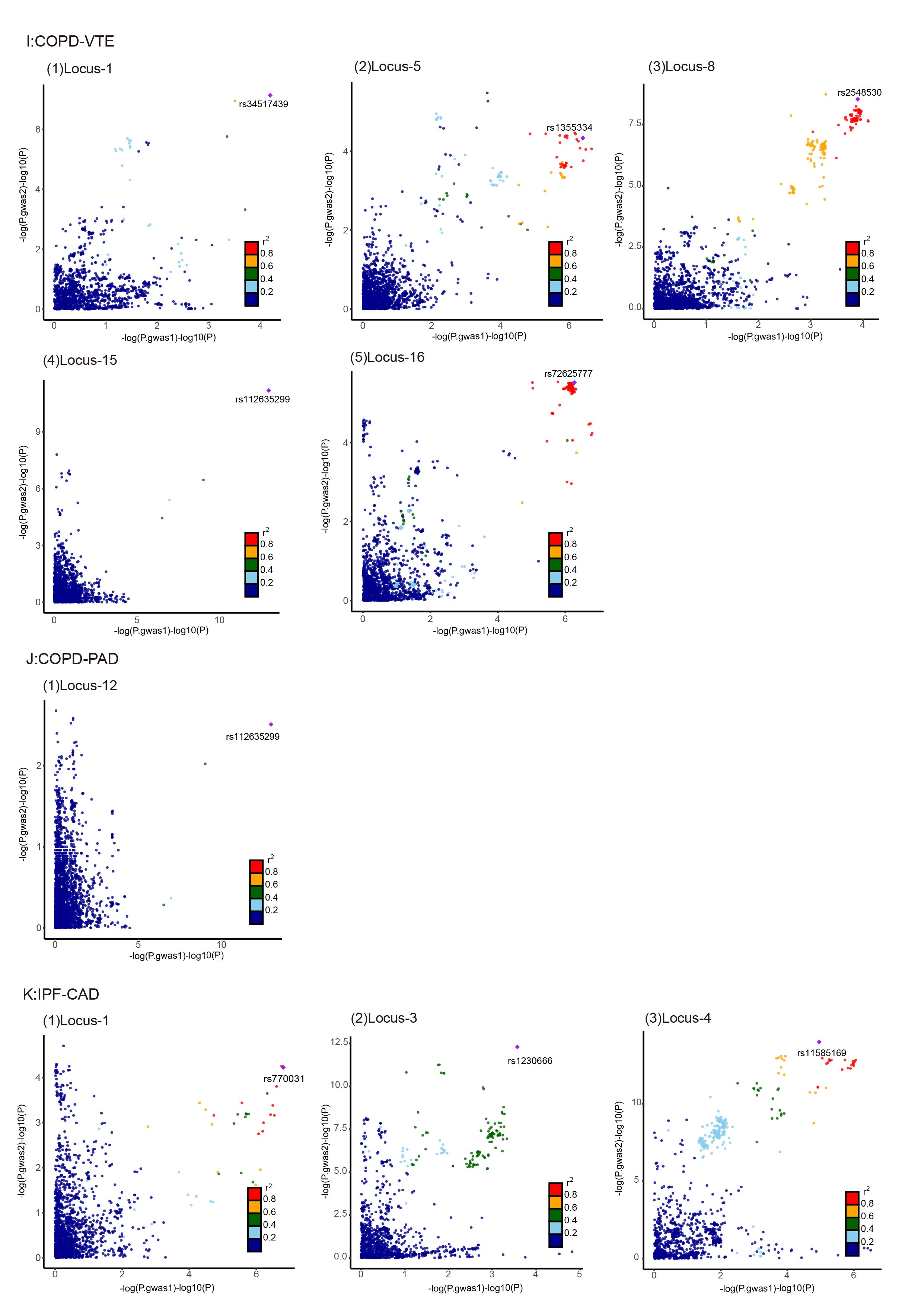

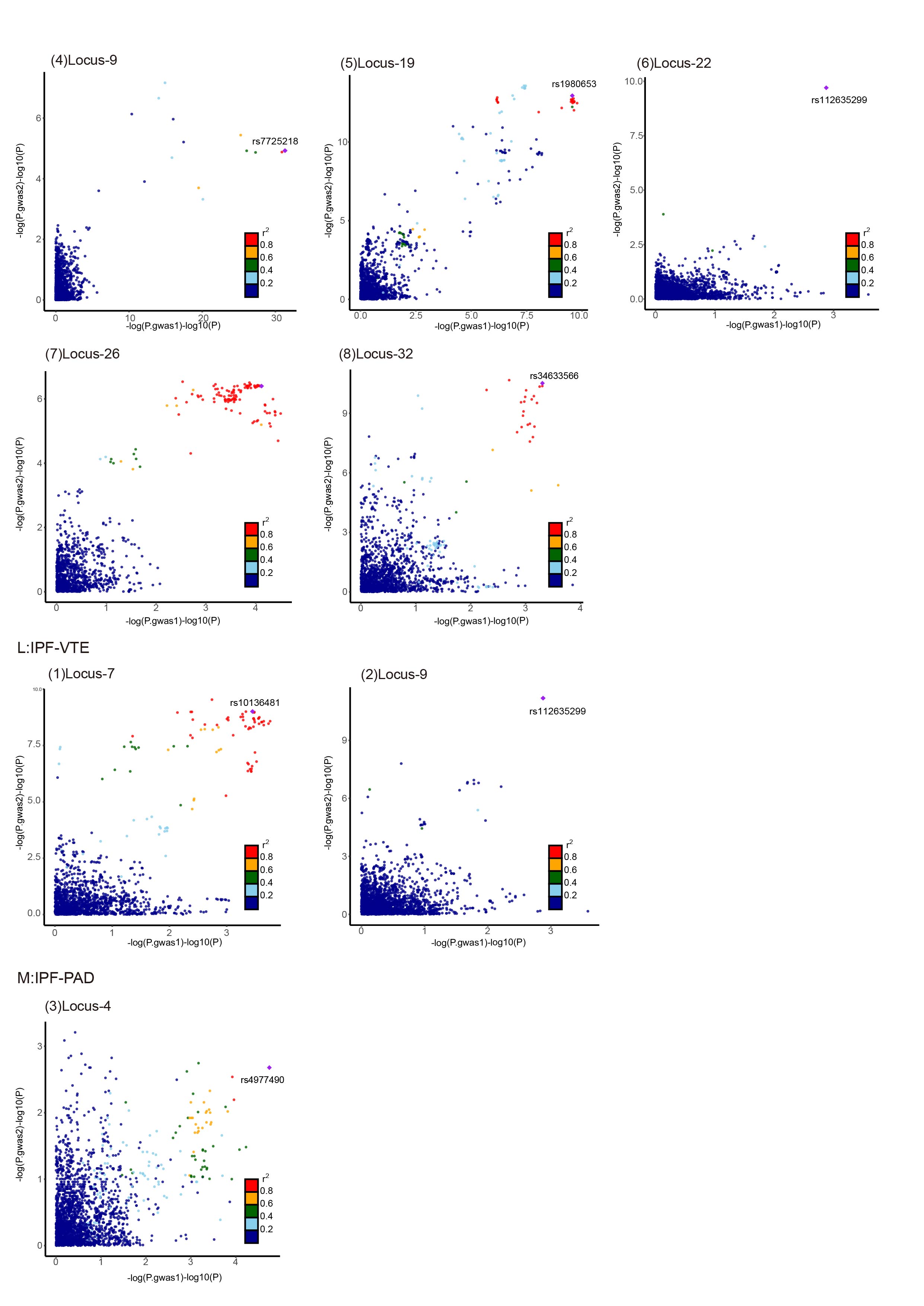

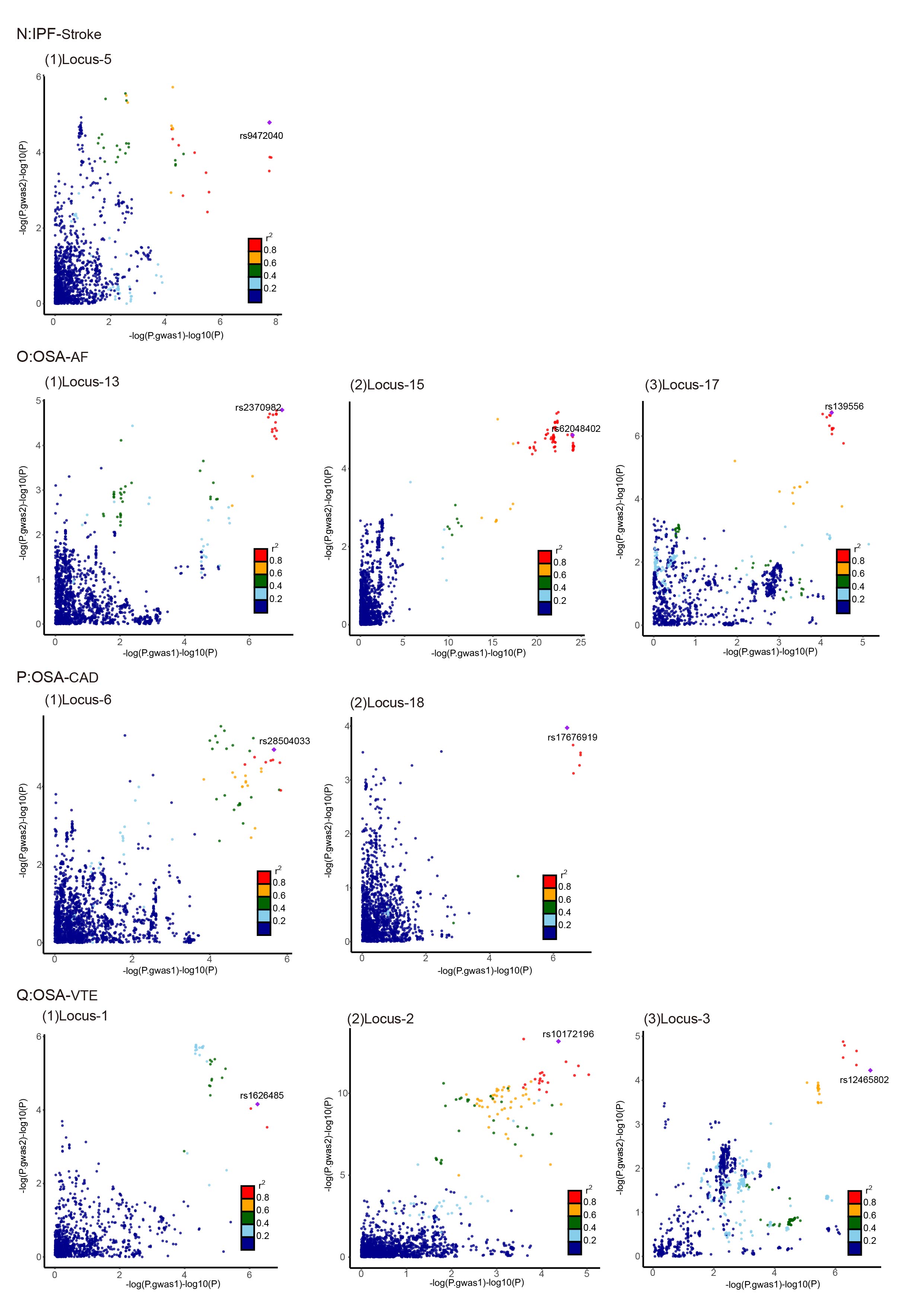

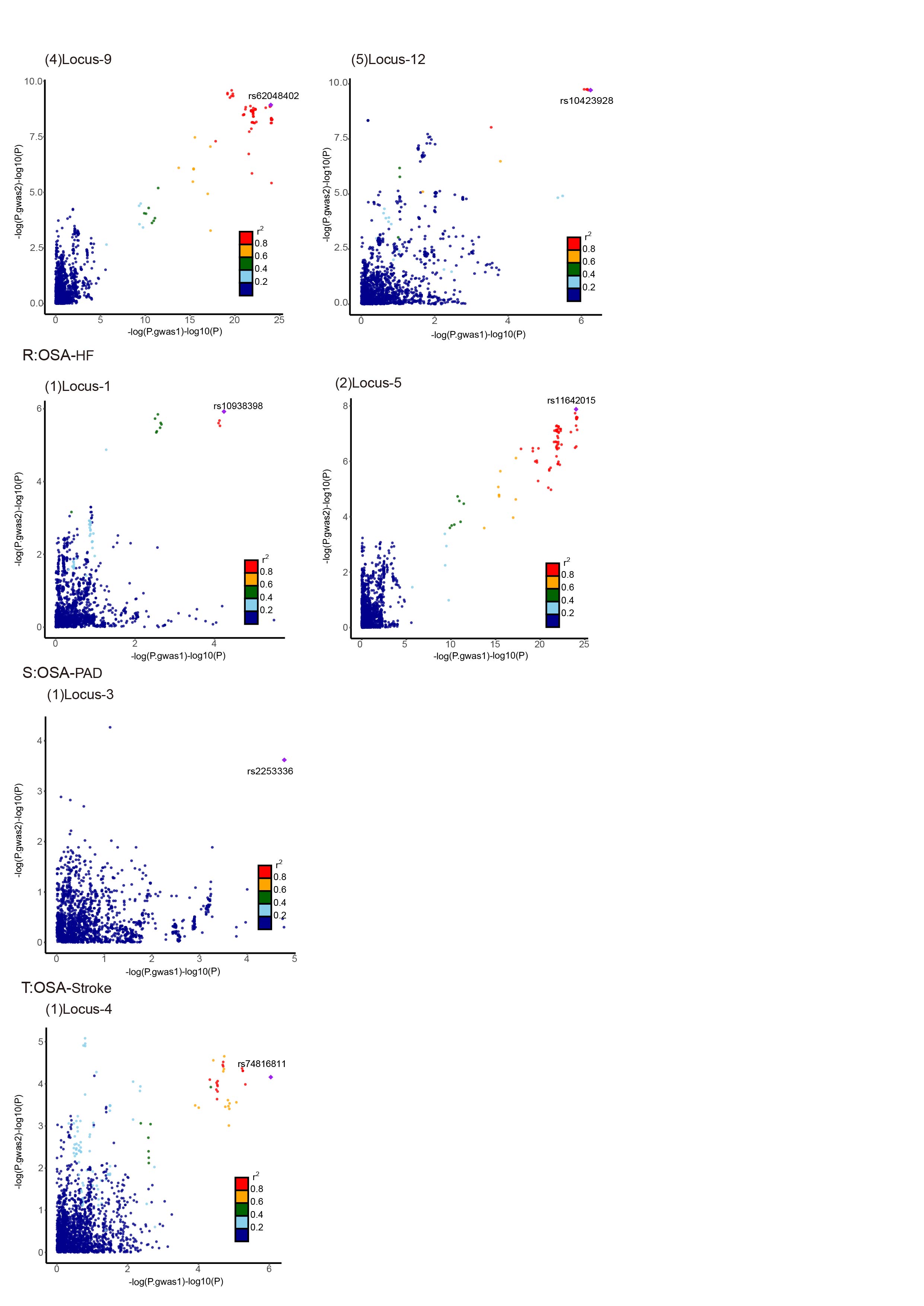

**Supplementary Fig. 4. LocusCompare Plots of 73 Colocalized Loci**

The LocusCompare plots of the genome-wide significant loci in the colocalization analysis, each dot represents a variant, the x-axis shows the −log10 PGWAS from the corresponding GWAS of respiratory diseases, and the y-axis shows −log10 PGWAS from the corresponding GWAS cardiovascular diseases. Each dot represented a genetic variant, with the candidate causal variant identified by pairwise colocalization analysis shown as a purple diamond. The color of other variants indicated their LD relationship with the candidate shared causal variant with the purple diamond from blue to red based on the 1000 Genomes Project European reference panel.

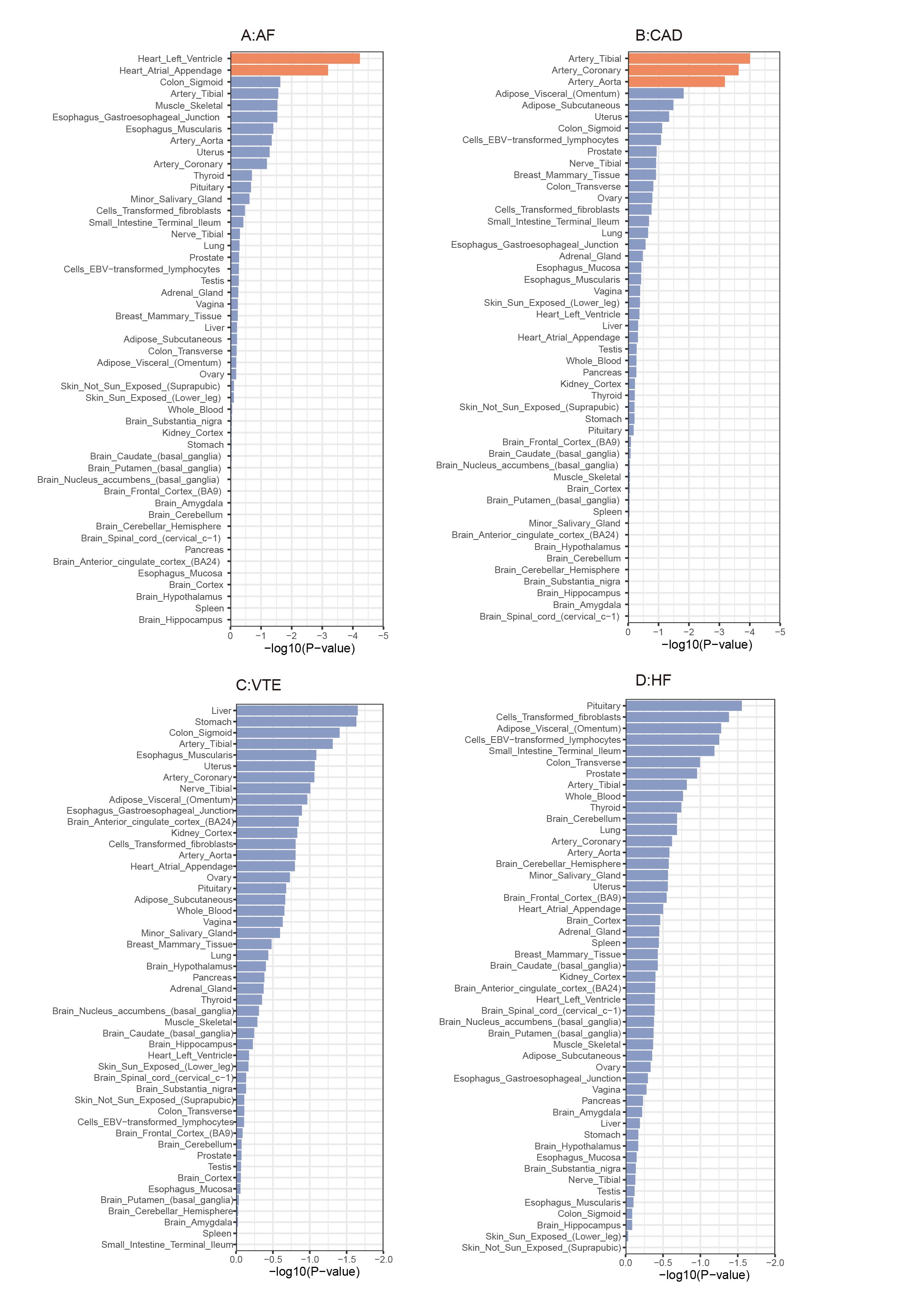

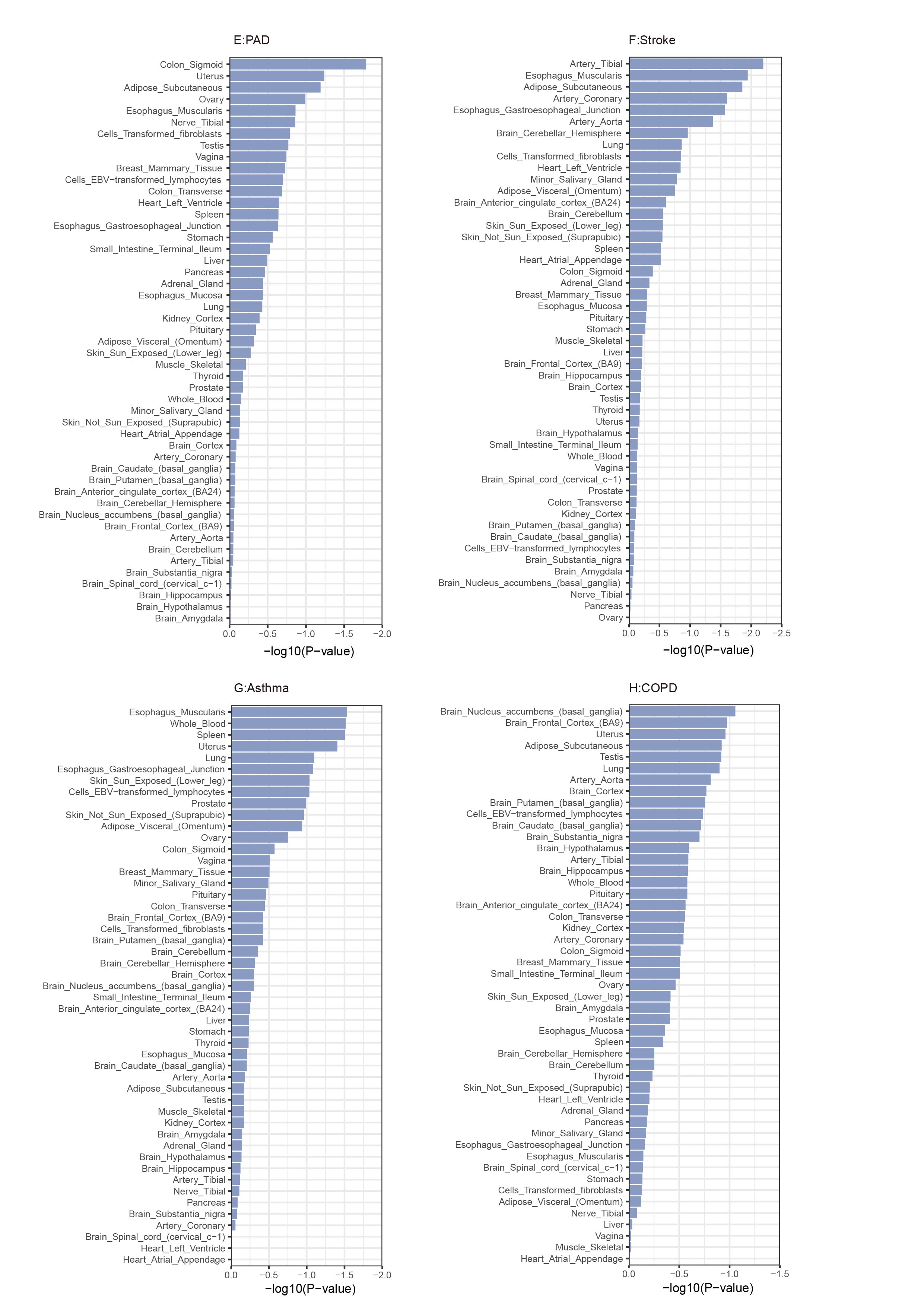

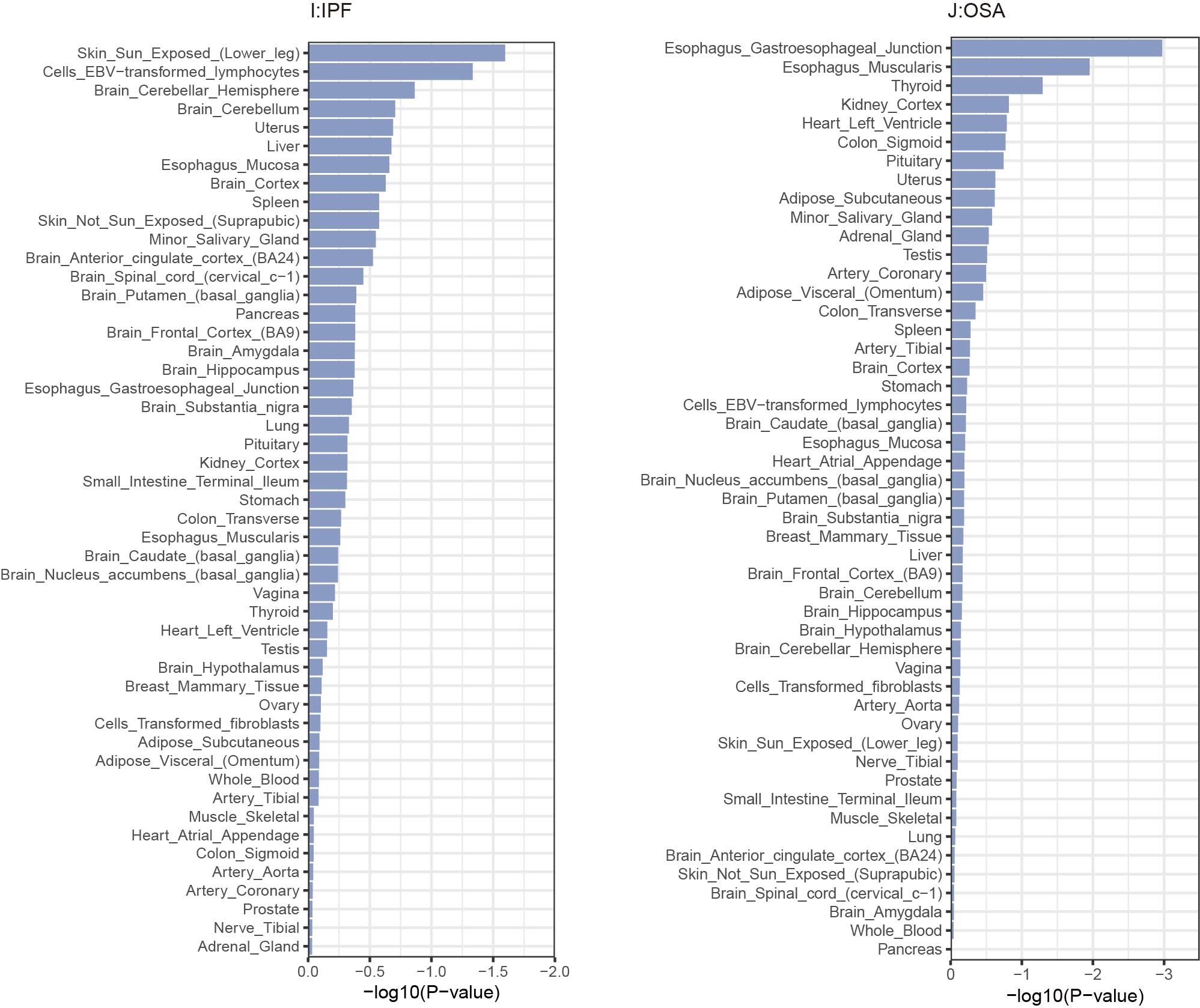

**Supplementary Fig. 5. The results of multiple-tissue analysis using gene expression data for the 24 trait pairs.**

Each point represents a tissue from the Genotype-Tissue Expression (GTEx) dataset. The orange color indicates a statistically significant p-value after conducting Bonferroni correction for multiple testing, which was < 1.02×10−3 (0.05/49) at -log10 (P) = 2.99.

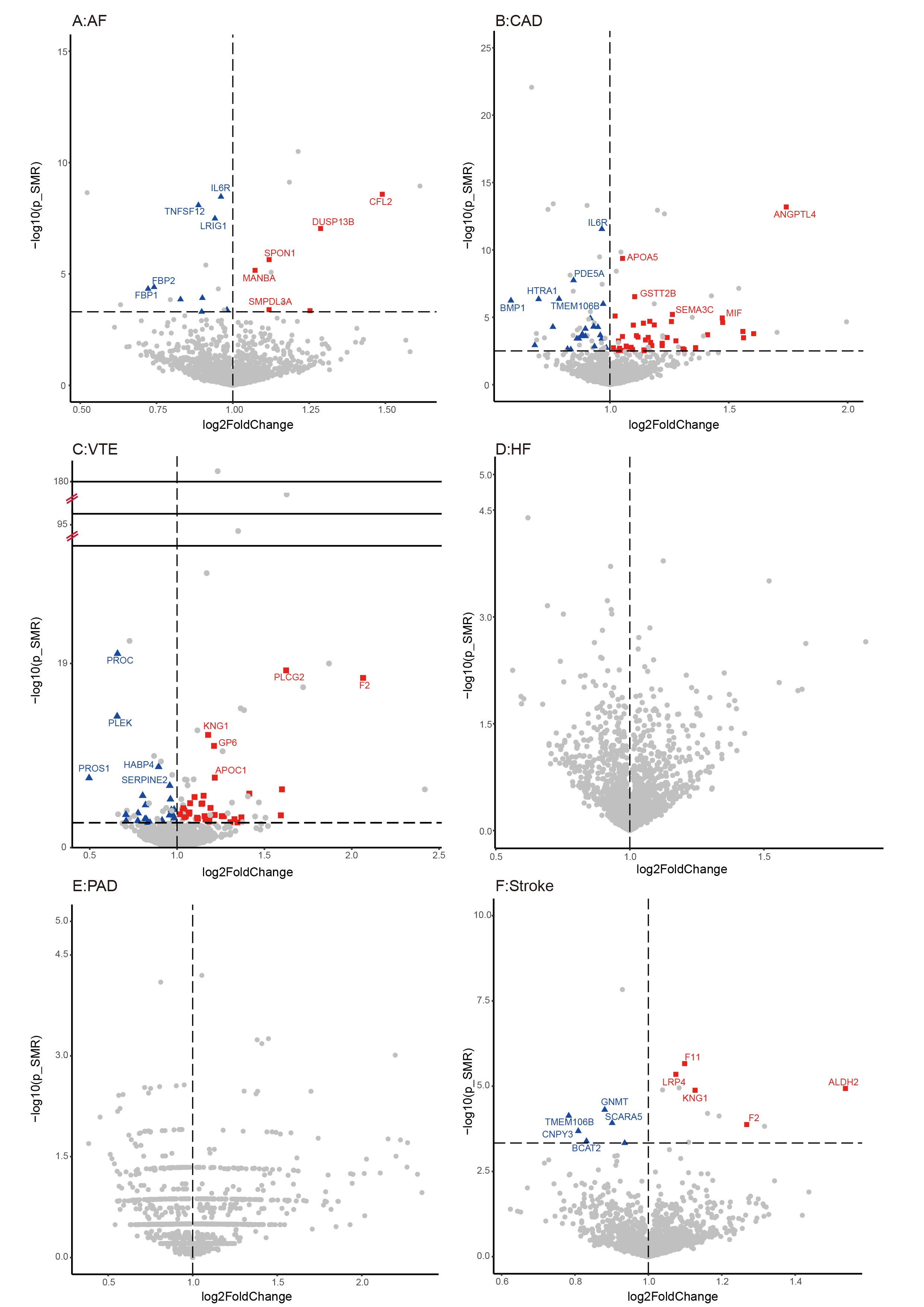

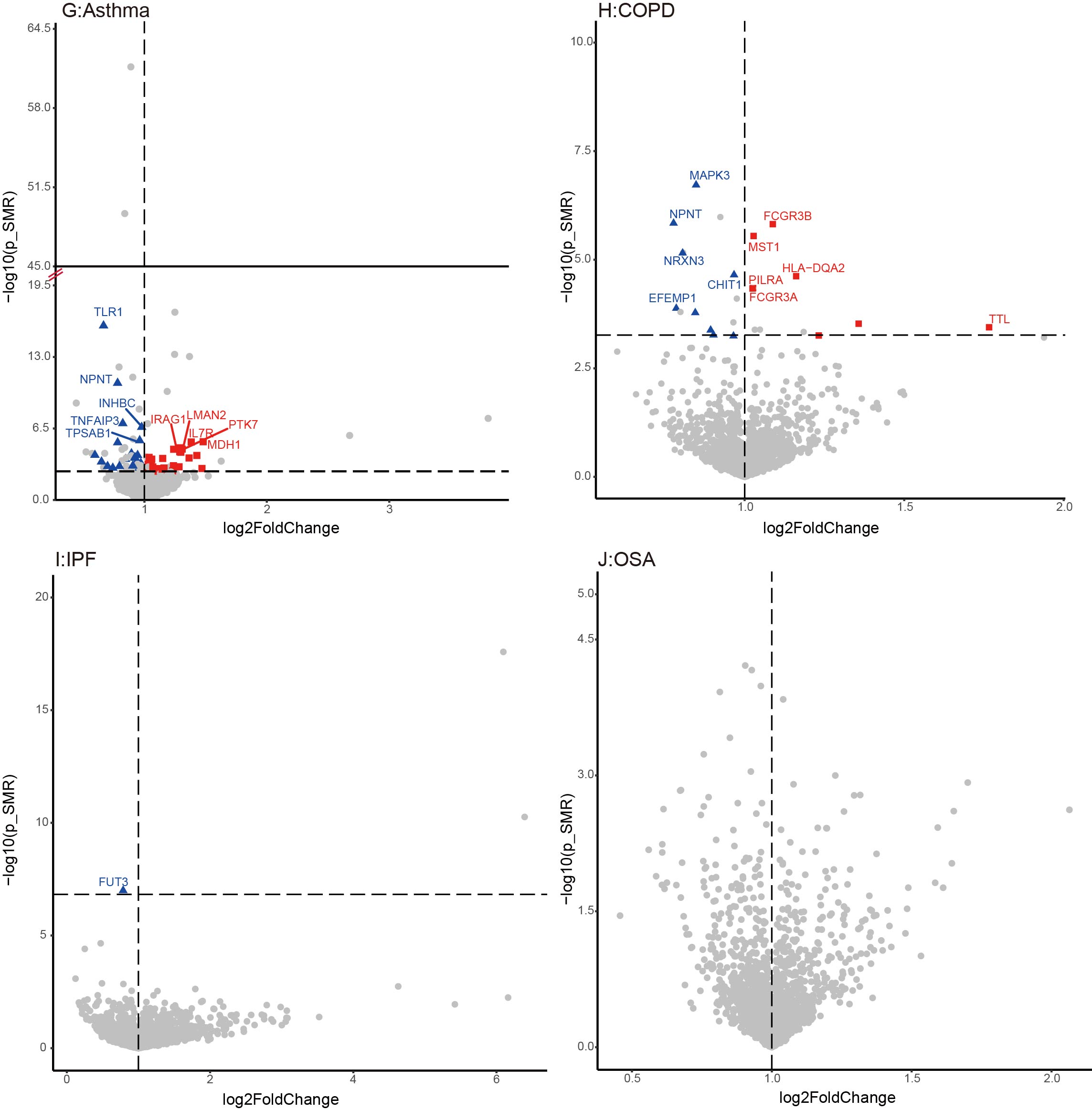

**Supplementary Fig. 6. Proteome-wide MR of the 4 respiratory diseases and 6 cardiovascular diseases.**

Volcano plot of associations between 1,708 proteins and each of cardiovascular diseases. Association strength between proteins and cardiovascular diseases according to their effect. Annotated proteins are the top five most significant that passed the threshold of proteome-wide MR and colocalization analysis. The red and blue colors represent positive and negative effects, respectively.

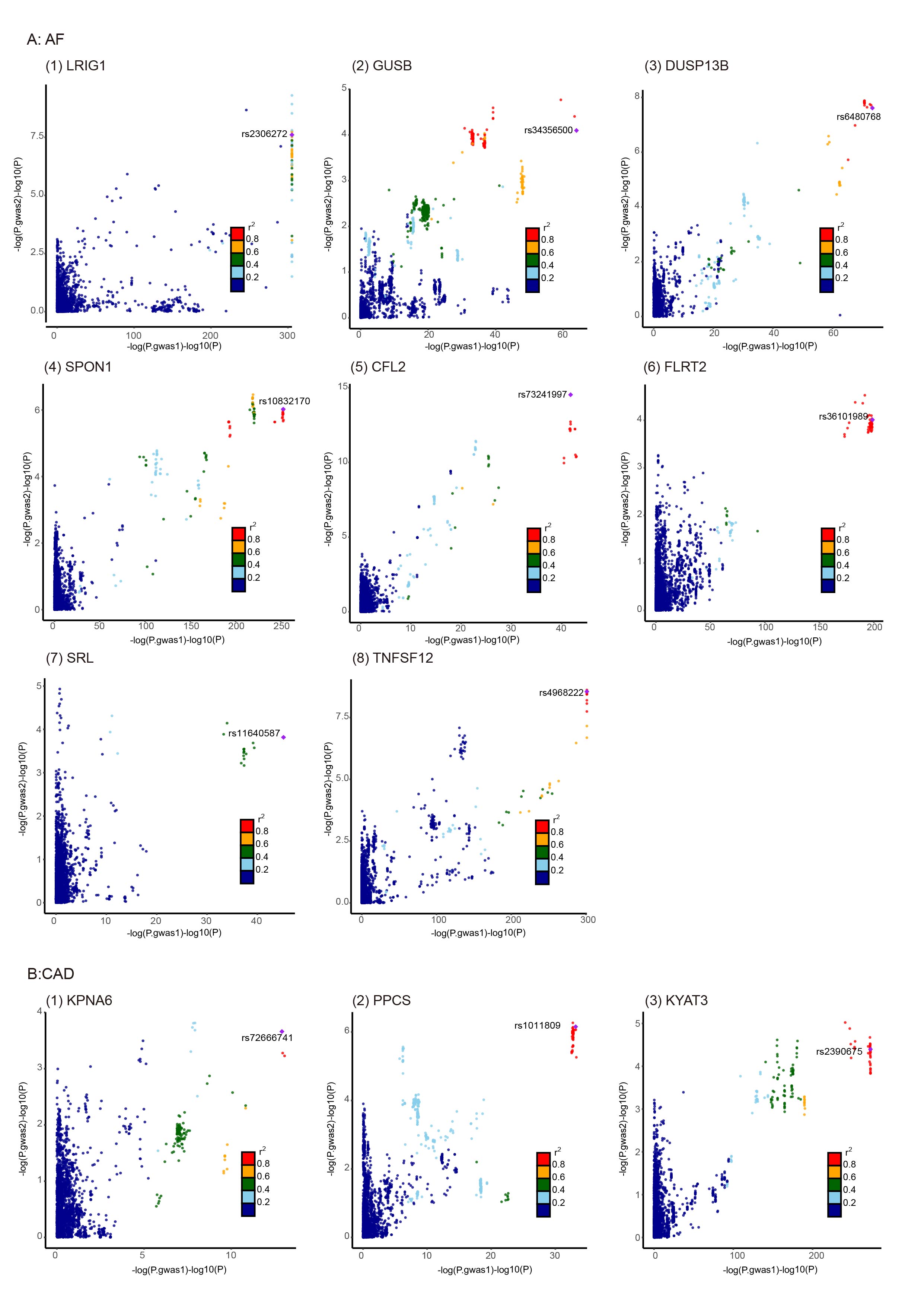

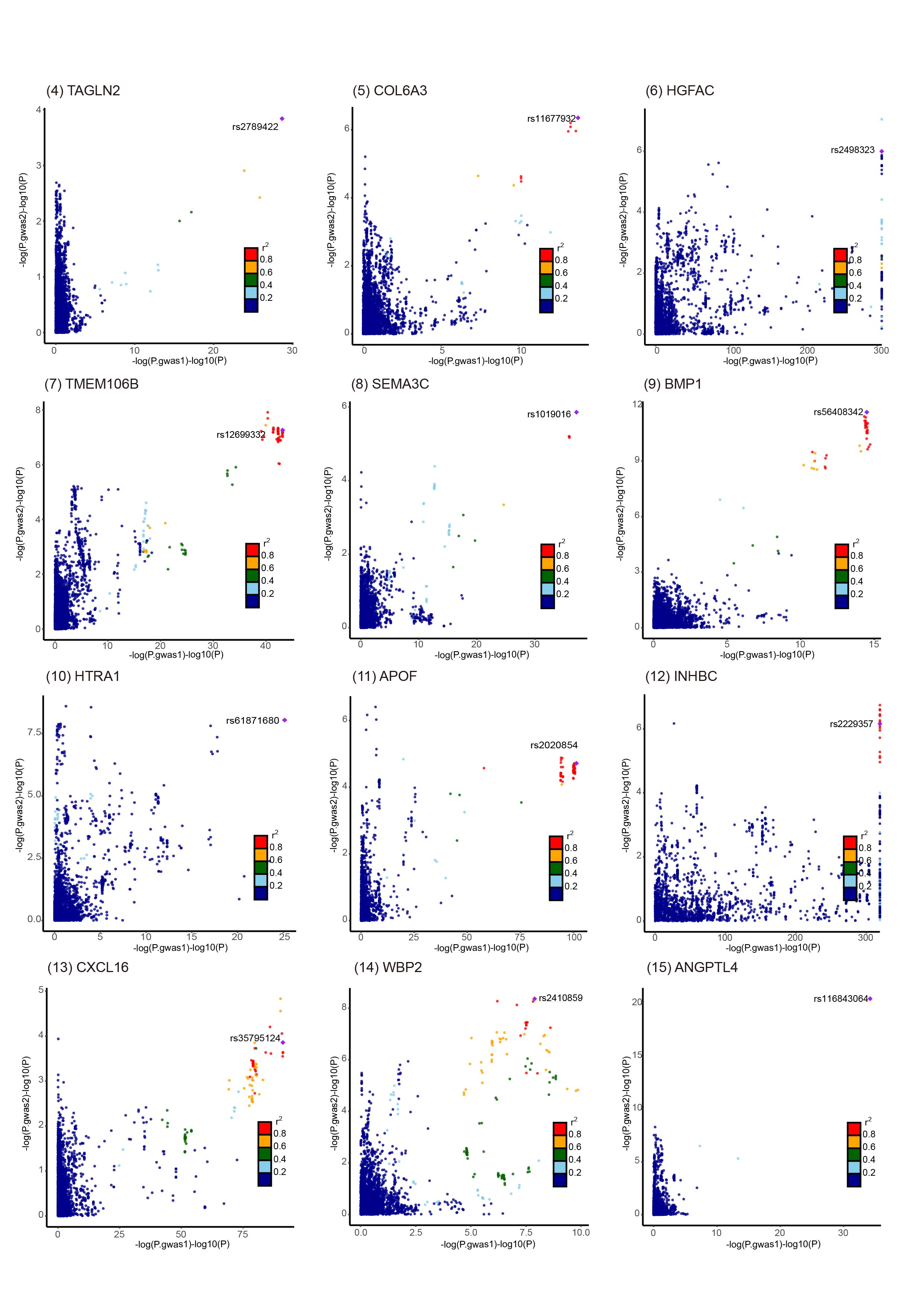

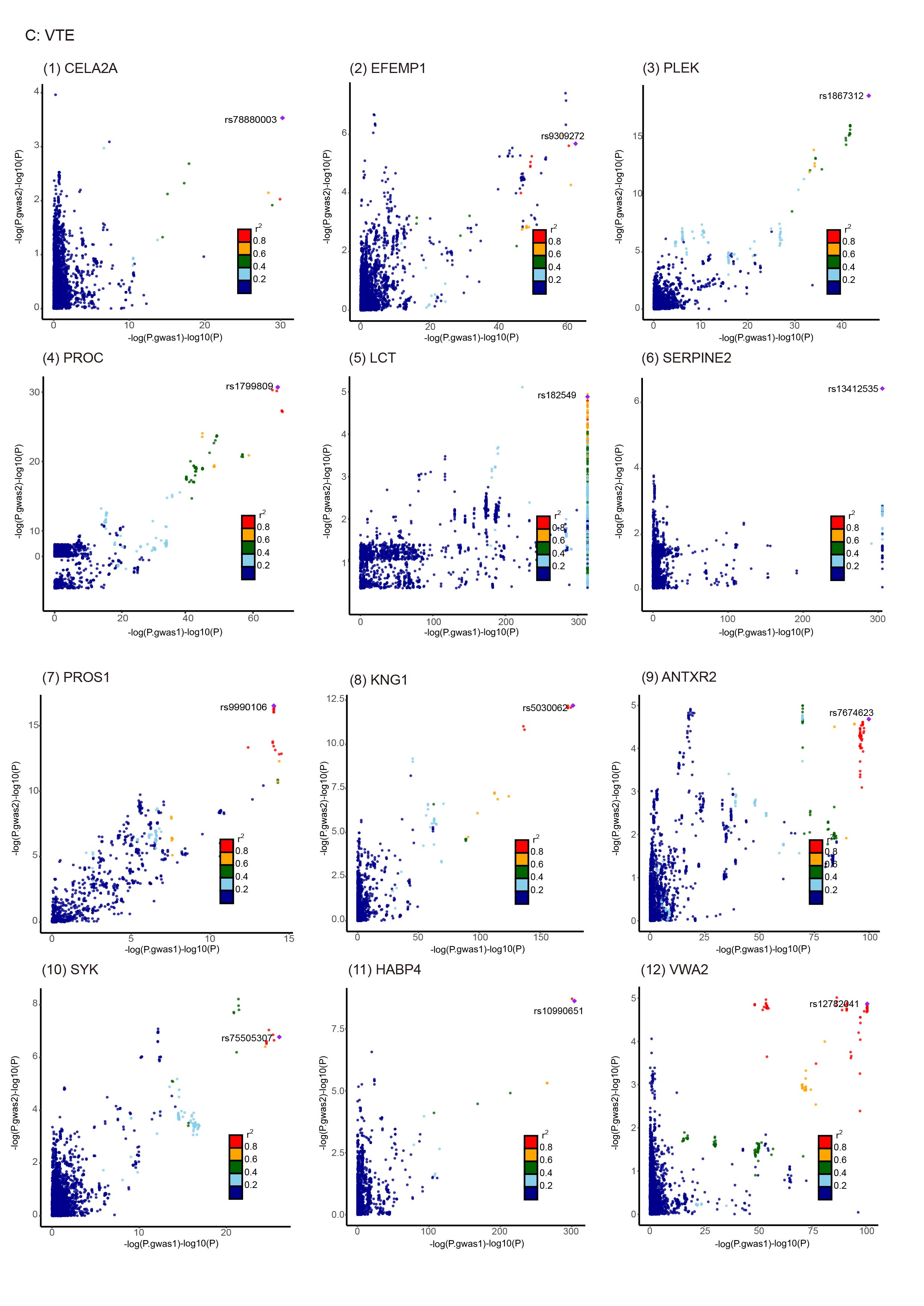

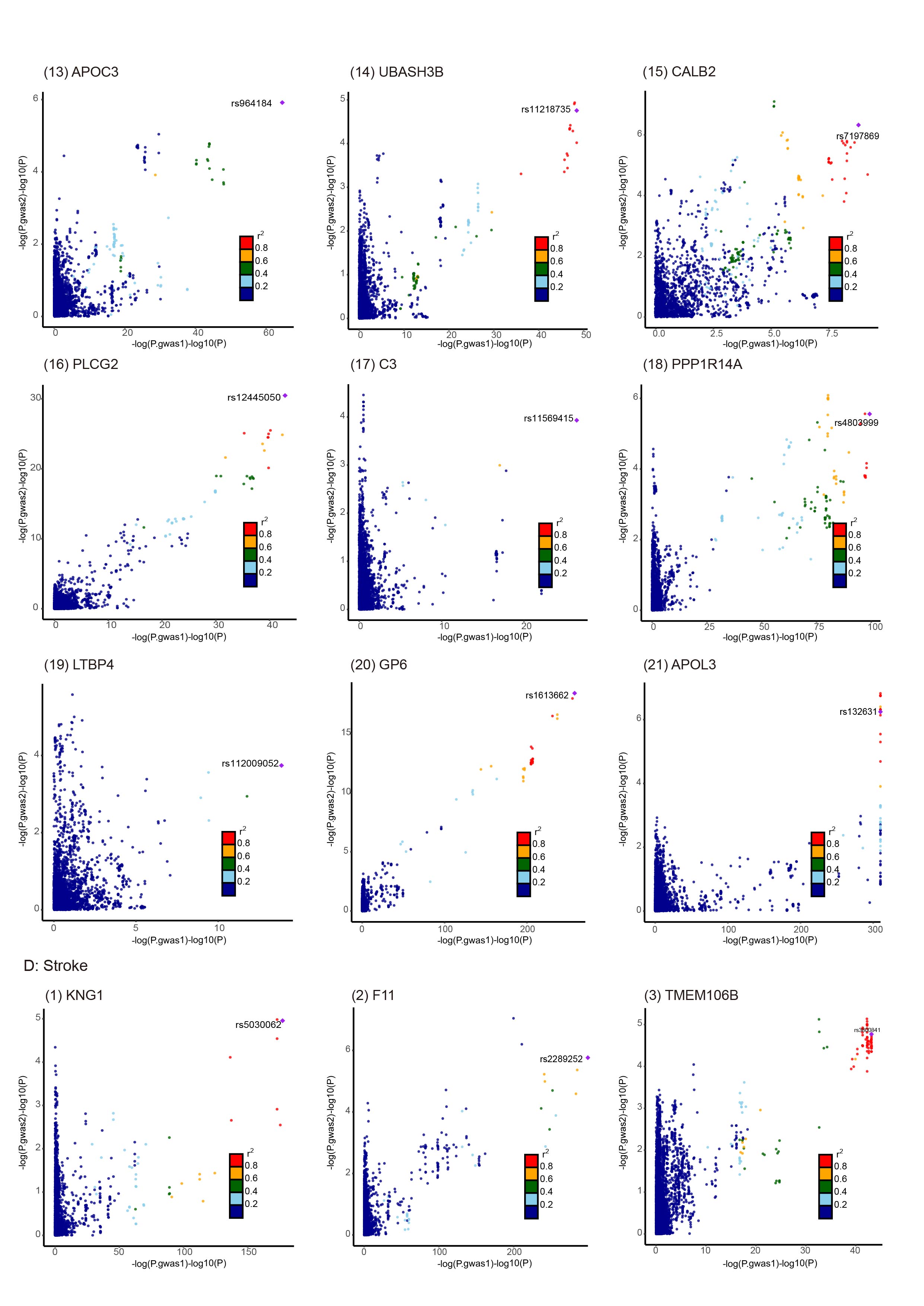

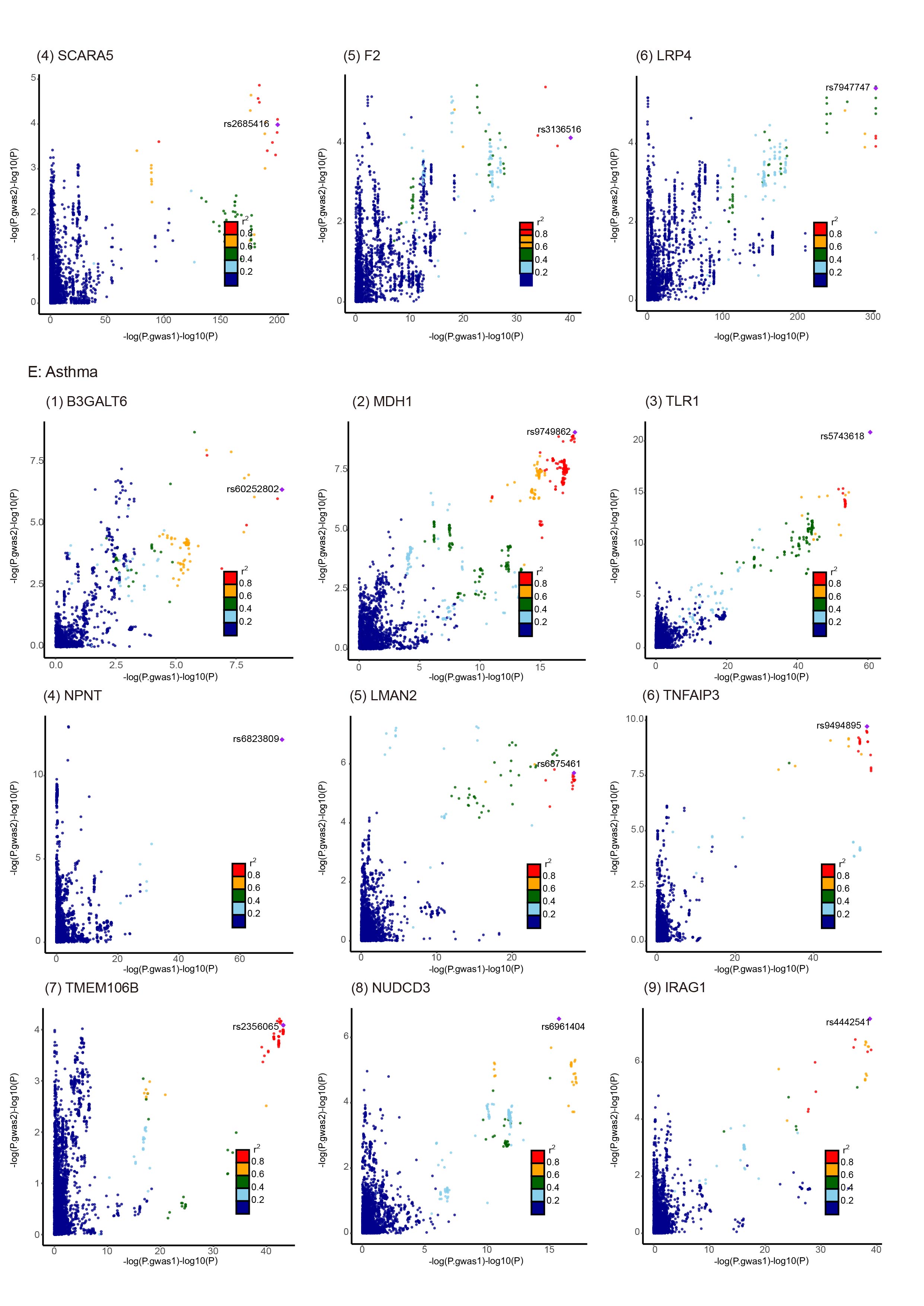

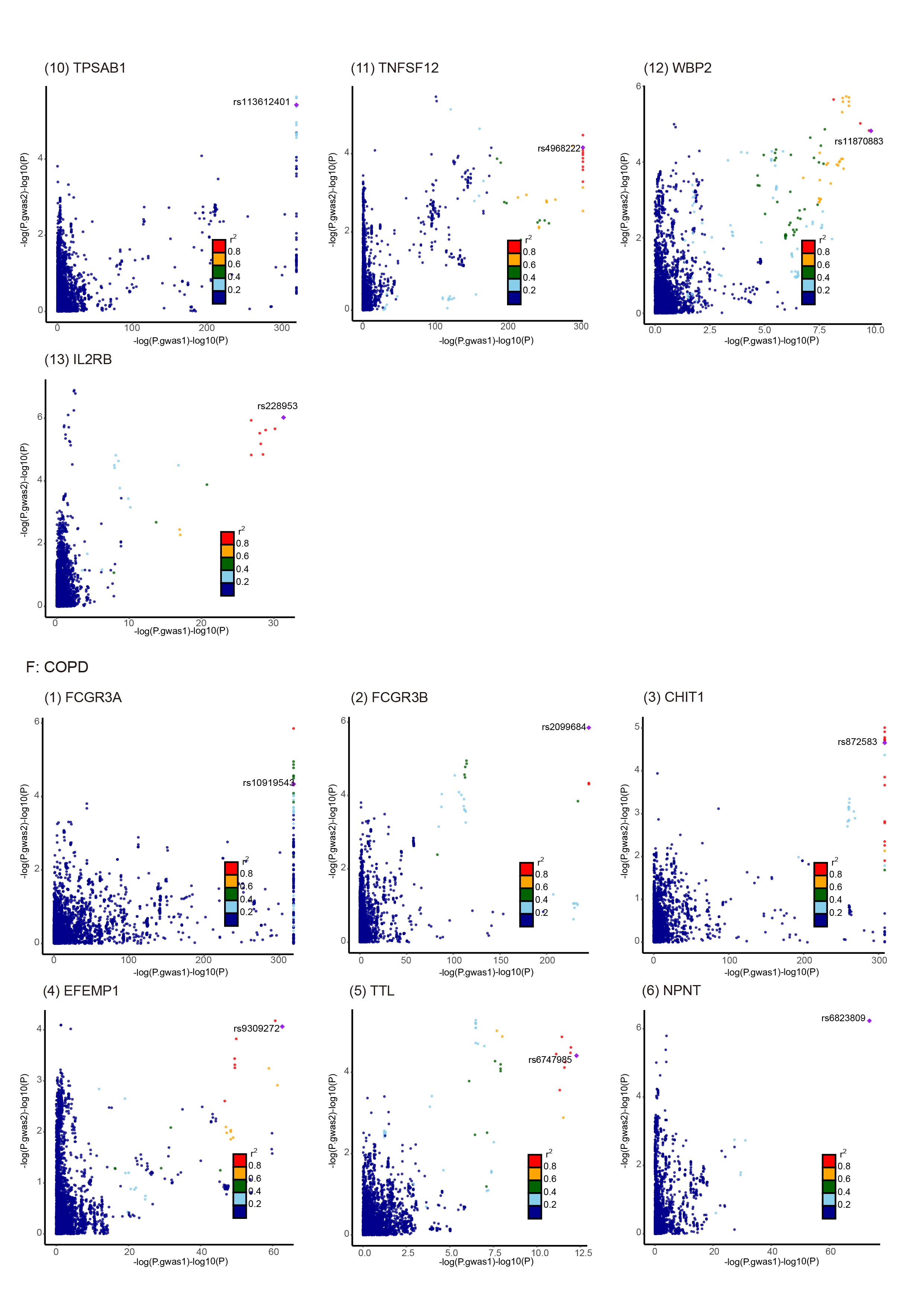

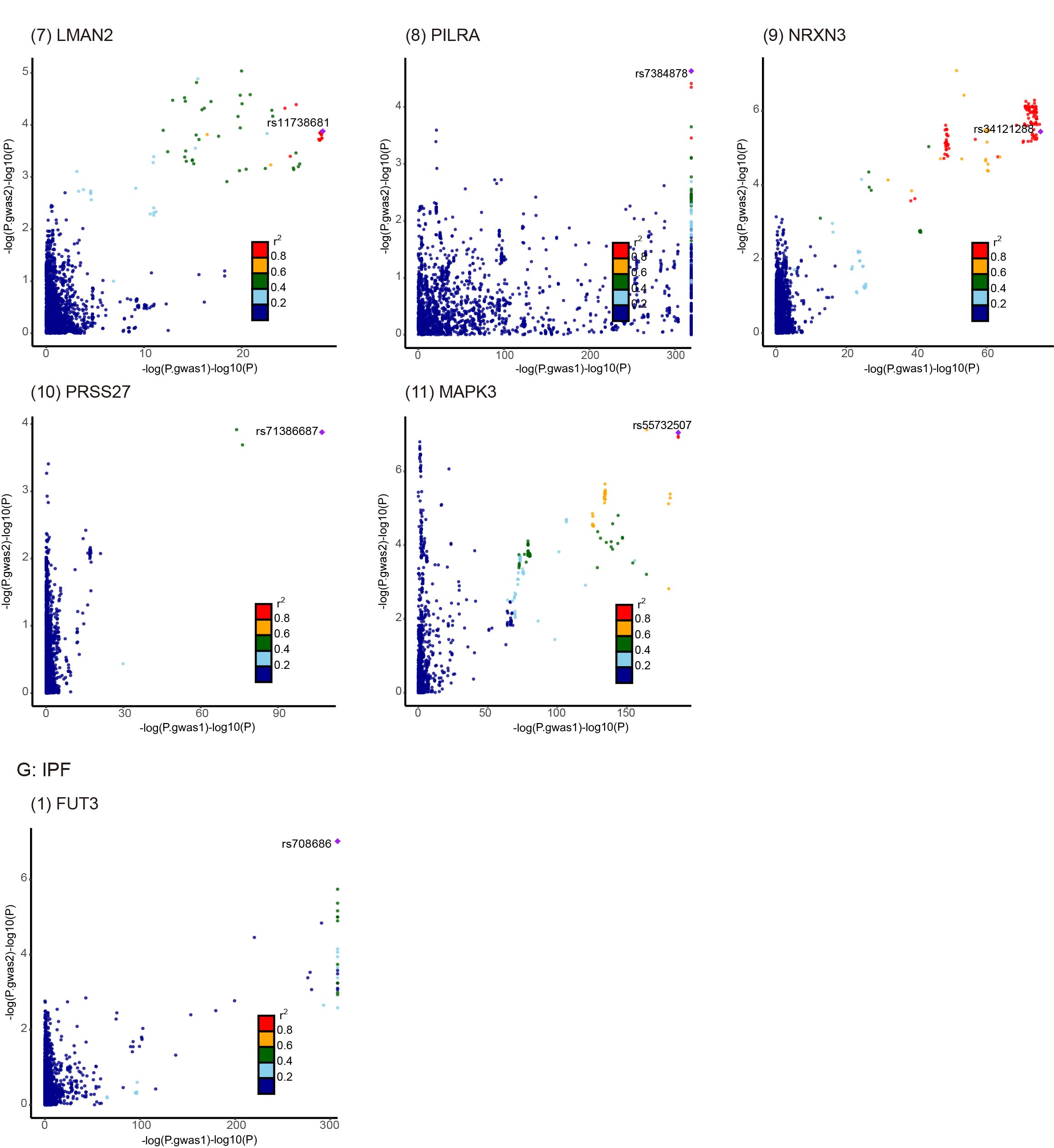

**Supplementary Fig. 7.** **The LocusCompare Plots plot for associations of causal plasma protein levels with respiratory diseases and cardiovascular diseases in colocalization analysis.**

Each plot on the plot represents a causal plasma protein, where the SNP with the most significant P value for cardiovascular disease is marked. The SNPs are color-coded based on linkage disequilibrium (r2) in Europeans with the lead variant, where those SNPs without available information are shown as dark blue dots. The X-axis of the panel shows the -log10 P values for associations with diseases, while the Y-axis shows the -log10 P values for associations with plasma protein levels.
